# Evaluating Prognostic Bias of Critical Illness Severity Scores Based on Age, Gender, and Primary Language in the USA: A Retrospective Multicenter Study

**DOI:** 10.1101/2022.08.01.22277736

**Authors:** Xiaoli Liu, Max Shen, Margaret Lie, Zhongheng Zhang, Deyu Li, Chao Liu, Roger Mark, Zhengbo Zhang, Leo Anthony Celi

## Abstract

**Background:** Although severity scoring systems are used to support decision making and assess ICU performance, the likelihood of bias based on age, gender, and primary language has not been studied. We aimed to identify the potential bias of them such as Sequential Organ Failure Assessment (SOFA) and Acute Physiology and Chronic Health Evaluation IVa (APACHE IVa) by evaluating hospital mortality across subgroups divided by age, gender, and primary language via two large intensive care unit (ICU) databases.

**Methods:** This multicenter, retrospective study was conducted using data from the Medical Information Mart for Intensive Care (MIMIC, 2001-2019) database and the electronic ICU Collaborative Research Database (eICU-CRD, 2014-2015). SOFA and APACHE IVa scores were obtained from the first 24 hours of ICU admission. Hospital mortality was the primary outcome. Patients were stratified by age (16-44, 45-64, 64-79, and 80-), gender (female and male), and primary language (English and non-English) then assessed for discrimination and calibration in all subgroups. To evaluate for discrimination, the area under receiver operating characteristic (AUROC) curve and area under precision-recall curve (AUPRC) were used. Standardized mortality ratio (SMR) and calibration belt plot were used to evaluate calibration.

**Findings:** A total of 173,930 patient encounters (78,550 MIMIC and 95,380 eICU-CRD) were studied. Measurements of discrimination performed best for the youngest age ranges and worsened with increasing age (AUROC ranging from 0.812 to 0.673 for SOFA and 0.882 to 0.754 for APACHE IVa, p <0.001). There was a significant difference in discrimination between male and female patients, with female patients performing worse. With MIMIC data, patients whose primary language was not English performed worse than English speaking patients (AUROC ranging 0.771 to 0.709 [p <0.001] for SOFA). Measurements of calibration applied to SOFA showed a statistically significant overestimation of mortality in the youngest patients (SMR 0.55-0.6) and underestimation of mortality in the oldest patients (SMR 1.54-1.57). When using SOFA, mortality is overestimated for male patients (SMR 0.92-0.97) and underestimated for female patients (SMR 1.05-1.11) while mortality is overestimated for English-speaking patients (SMR 0.85) and greatly underestimated for non-English speaking patients (SMR 1.4). In contrast, the calibration applied to APACHE-IVa shows underestimation of mortality for all age groups and genders.

**Interpretation:** The differences in discrimination and calibration with increasing age, female gender, and non-English speaking patients suggest that illness severity scores are prone to bias in their mortality predictions. Caution must be taken when using these illness severity scores for quality benchmarking across ICUs and decision-making for practices among a diverse population.

**Funding:** Z.B.Z was funded by the National Natural Science Foundation of China (62171471).

**Research in context:** *Evidence before this study:* We searched PubMed, arXiv, and medRxiv from the inception of the database to July 10, 2022, for articles published without language restrictions. The search terms were (illness severity score OR SOFA OR APACHE-II OR APACHE-IV OR SAPS) AND (evaluation OR performance OR bias) AND ((age OR older OR elderly OR 65 years old OR 80 years old OR subgroup) OR (gender OR Female OR male) OR (language speaking OR English speaking)). Multiple studies have explored the performance among their concerned subgroups with limited patients and hospitals such as over 80, older with sepsis, and surgical patients. Although a small number of studies have presented the performance of scores by age groups, they have not systematically examined the differences and bias between younger and older patients in depth. Few articles analyzed the differences between men and women. No study has discussed the evaluation performance between Non-English and English speakers. We identified that no studies have comprehensively reported the potential bias of clinical scores in the assessment of subgroups classified by age, gender, and English-speaking.

*Added value of this study:* To our best knowledge, we are the first to conduct a systematic bias analysis of the SOFA and APACHE-IVa scores to assess in-hospital outcomes across age (16-44, 45-64, 65-79, and 80-), gender (male and female), and English speaking (Yes and No) subgroups using multicenter data from 189 U.S. hospitals and 173,930 patients episodes. The assessment was performed covering discrimination (AUROC and AUPRC) and calibration (SMR and Calibration belt plot). We found that the AUROCs between the two scores decreased significantly with age. The illness severity exists underestimation for oldest patients and serious overestimation for youngest patients using SOFA score. Both scores demonstrated slightly better AUROCs for males. For Non-English speaking patients, SOFA showed a large reduction in AUROC and very significant underestimation compared to English speakers. Furthermore, there exists higher observed mortality of older patients, females, and Non-English speakers compared to their respective other subgroups using the same SOFA score.

*Implications of all the available evidence:* The aging of the ICU, especially the extremely rapid growth of patients over 80 years old. They exhibit unique characteristics with more comorbidities, frailty, worse prognosis, and the need for more humanistic care, which has evolved into a serious challenge for early clinical triage, diagnosis, and treatment. Females are more likely to withhold pain and not be transferred to the ICU for treatment, which leads to potentially more critical severity illnesses admitted to ICU compared to males. SOFA and APACHE-IVa scores are very important basis and standards for early ICU assessment of illness severity and decision-making. While these general phenomena were noticed in clinical practice of the mentioned subgroups, there is a lack of clear and detailed quantitative analysis of the bias in the use of these scores to protect these vulnerable populations and prevent potential unintentional harm to them. The U.S. is a multicultural and racially integrated country, and the number of Non-English speakers is rising every year which reflects greater socioeconomic and ethnic disparities. Limited communication can also have an impact on patient assessment and treatment. However, the use of the SOFA score for the evaluation of this group of patients has not been reported to date. In this study, we used multicenter data with a large sample size to identify potential bias using the SOFA and APACHE-IVa scores for all mentioned special groups of patients.

## Introduction

Illness severity scoring systems such as the Sequential Organ Failure Assessment (SOFA) and Acute Physiology and Chronic Health Evaluation IVa (APACHE IVa) are used to predict prognosis among patients in the Intensive Care Unit (ICU)s [1-5]. The scores provide benchmarks to predict outcomes, triage patients, support decision-making, assess ICU performance, and allocate scarce resources [6-9]. Despite the need for fair, unbiased systems, these illness severity scores are limited by population-level prognostic estimation, leading to variable performance across subgroups such as ethnicity [8,9,15,16]. Additional sources of bias could be found in subgroups categorized by age, gender, and English speaking, but bias in these subgroups has not been fully explored [17-20].

Age plays a significant role in ICU care, and especially patients over 80-years-old demonstrate the fastest population growth rate in the critical care setting [17,21-23]. According to Daniele et al., an illness severity score called the Simplified Acute Physiology Score (SAPS) III was prone to underestimate the risk of death for patients older than 80-years-old when compared to patients younger than 40-years-old [27]. And a study conducted by Fernando et al. further indicated that the accuracy of the SAPS III score could be improved by accounting for performance status and comorbidity for older patients [28]. In relation to gender bias, women are often subject to inequality such as delays in treatment [18]. In the ICU, women face a higher risk of mortality. Recently, Atanas T et al. performed an observation analysis on gender differences containing 45,0948 patients (77,803 admitted to the ICU), and there exists a significantly higher risk of mortality in women when evaluated by SAPSII score (odds ratio 1.008 [1.004–1.012]) [18]. A significant portion of the population has limited English proficiency (LEP) in the U.S, and this population is growing every year [19,24]. A recent study reported their LEP rate to be 11% in an academic medical center [25]. However, the impact of LEP on the quality of healthcare delivery has received little attention [25,26,44]. A newly published research analyzed the effect of LEP on mortality in 8974 patients with sepsis. They reported LEP was a significant predictor of death and a higher qualifying SOFA score (total score when SOFA firstly larger than 2 within two days) of LEP patients was observed [29].

Although a small number of studies have explored the characteristics of these special groups, the sample size and source (multicenter) of the studies are limited. The comprehensive bias evaluation of clinical scores in assessing these groups has not been the focus of these studies. In addition, the use coverage of the SAPS score is much lower than that of SOFA and APACHE scores [1,2,7-10]. In this multicenter retrospective study, we sought to evaluate the existing bias of SOFA and APACHE-IVa scores in the multiple groups divided by age (16-44, 45-64, 65-79, and 80-years old), gender (female and male), and language speaking (English and Non-English). The discrimination and calibration of these predictive scoring systems with ICU patients’ hospital mortality were our primary concern performances.

## Methods

### Data sources

Two high-quality clinical databases were utilized for this study. The Medical Information Mart for Intensive Care (MIMIC) is an open shared clinical database containing ICU admission data at a tertiary academic hospital, the Beth Israel Deaconess Medical Center (BIDMC), in Boston, Massachusetts [30-32]. Our analysis included both MIMIC-IV with data from 2008 to 2019, as well as MIMIC-III with data from 2001 to 2012, with duplicate data included in both versions used only once. Altogether, MIMIC contains 83,478 patients with 104,843 admissions and 114,316 ICU admissions. The eICU Collaborative Research Database (eICU-CRD) is generated from the Philips telehealth system covering 208 hospitals and 9.13% of teaching hospitals across the U.S [15,33]. It contains 139,367 patients, 166,355 admissions, and 200,859 ICU admissions from 2014 to 2015 in the latest release of V.2.

From the two databases, we excluded episodes where the patient was younger than 16 years old or ICU length of stay is less than 24 hours. We further excluded patient episodes without APACHE-IVa score in the eICU-CRD, as well as encounters where SOFA scores could not be calculated from both databases. Specific data extracted from MIMIC include but are not limited to demographic information such as (age, gender, race, ethnicity, language speaking, admission type, range of admission year, discharge status), clinical data including laboratory measurements, vital signs, and treatment records, as well as diagnosis codes for billing, etc. The eICU-CRD has similar demographic and clinical data information records as MIMIC, though with a higher granularity of physiological measurements and the addition of calculated APACHE-IV(a) score for each admission [15]. To facilitate sub-group analysis, patient age is divided into 4 categories: 16-44, 45-64, 65-79, and over 80 years old. The patient’s primary language spoken was divided into English with clear identification and Non-English in the MIMIC.

The de-identification and anonymization were both strictly implemented in the MIMIC and eICU-CRD databases. Our retrospective study was exempted by the ethical review committee of the US.

### Statistical analysis

Hospital mortality was selected as our outcome of interest. For the SOFA score, as it is not originally intended for mortality prediction, we used two methods to characterize its relationship with mortality: 1) two logistic regression functions were fitted using their original continuous form from 0 to 24 points based on their respective datasets with 10% randomly selected patients [9]; 2) three risk categories (≤7, 8-11, and ≥12) previously proposed to facilitate triage and scarce resource allocation in COVID-19 pandemic [15,34]. For the APACHE-IVa score, the mortality prediction of each eICU patient episode was directly acquired from a database. To evaluate the statistical significance of age, gender, and language speaking, we separately performed the logistic regression analysis using scores and those variables in two study cohorts. Additionally, we conducted an extra calibration analysis of SOFA score, as in Sarkar et al, to assess performance in different severity levels with risk categories of 0-5%, 5-10%, 10-20%, 20-50%, and over 50% [15].

We conducted the discrimination analysis for each subgroup of age, gender, and primary language via the area under receiver operating characteristic(AUROC) and area under the precision-recall curve (AUPRC) with their associated 95% confidence interval (CI) were also reported. The AUROC was a normally used graphical plot of the true positive rate (TPR) against the false positive rate (FPR) [15,35]. And AUPRC was suitable for imbalanced data classification, a graphical plot of positive predictive value (precision, PPV) against the TPR [36]. We measured calibration using standardised mortality ratio (SMR) and Calibration Belt plot for both scores [37]. Descriptive statistics of patient characteristics were reported using median (25^th^, 75^th^) percentiles (IQR) or proportions. The significant differences between groups were compared using the student’s *t-test* or *X*^*2*^ test for categorical variables, and Wilcoxon rank-sum test or Kruskal-Wallis test for continuous variables, as appropriate. 200-fold bootstrap resampling iteration was used to calculate 95% CI, and 2-tailed *P* < 0.05 was used as a threshold for statistical significance. All statistical analyses were performed using Python version 3.8 (sklearn, pyroc, scipy, and tableone package), and R version 4.1.1 (ems, dplyr, forestplot, givitiR, gbm, and rsq package).

## Results

As shown in **Figure 1**, patients were separately extracted from both MIMIC and eICU-CRD datasets. MIMIC contains 104,422 separate hospital admissions at BIDMC across 19 years. 78,550 patient (75.2%) episodes with an 11.1% hospital mortality rate were included in this study. In eICU-CRD, 166,355 hospital admissions were drawn from 208 hospitals across the U.S, and 95,380 cases (57.3%) with a 9.0% hospital mortality rate were selected for this analysis. Altogether, our study consisted of 173,930 cases with 17,338 deaths in the final analysis.

**Figure 1.**
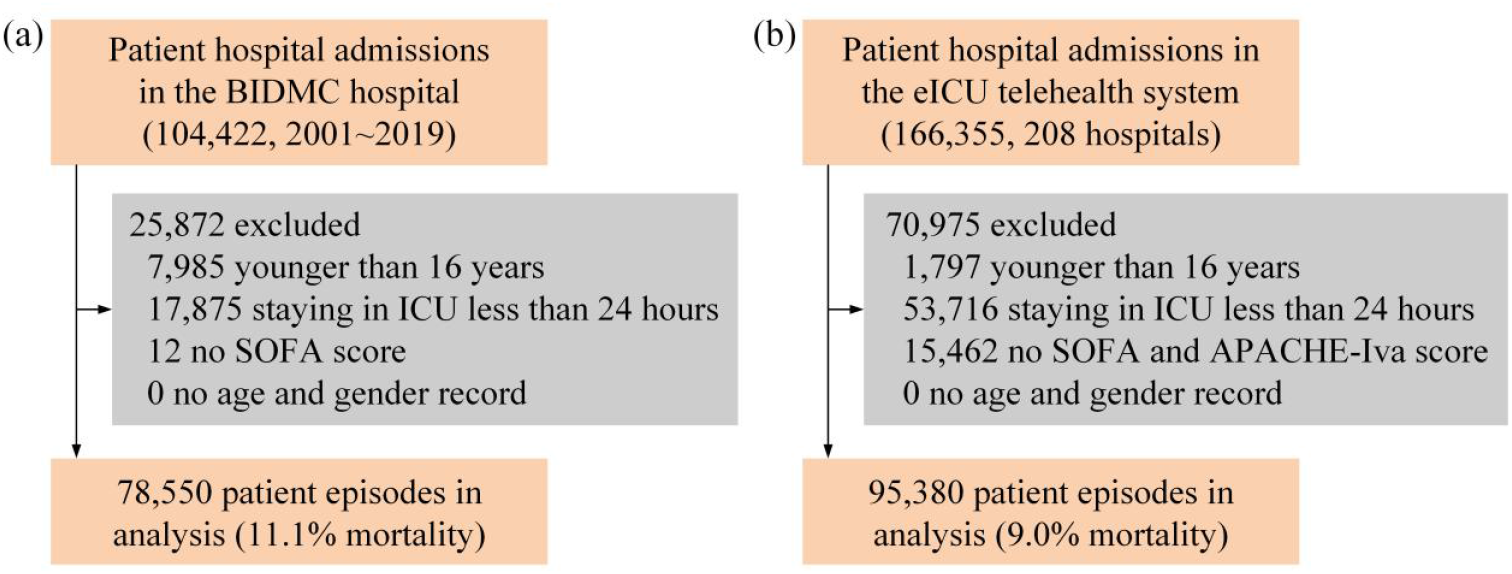
Study flow of the two study cohorts. (a) MIMIC, (b) eICU-CRD.

Baseline characteristics of the two study cohorts are shown in **Table 1**. In the MIMIC cohort, the median (IQR) age was 66 (54-78) years. Non-survivors were significantly older than survivors (66.0 yr vs. 73.0 yr), and the 16-44 yr age group has the lowest mortality compared with others (6.4% vs. 34.8% of 65-79 yr group, p<0.001). 43.6% of patients were female. BMIs of non-survivors were significantly lower (26.5 vs. 27.7, p<0.001). There is a higher percentage of non-English speakers among non-survivors when compared to survivors (36.7 vs. 26.8, p<0.001). 72.1% of English speakers and higher mortality with a significant difference for non-English speakers (14.6% vs. 9.7%). 68.6% of white and 25.7% of CCU admitted patients showed the largest percentages in ethnicity and first care unit. The higher SOFA scores for dead patients during the first ICU admission day were recorded (7.0 points vs. 4.0 points). The significantly higher percentage of comorbidity was measured by CCI score, higher ventilation receivers, and higher comfort therapy receivers of deaths than survivors (7.0 points vs. 5.0 points, 57.8% vs. 37.7%, 14.3% vs. 3.3%). The longer ICU duration of non-survivors (3.9 days vs. 2.3 days). In the eICU-CRD cohort, the significant difference between survivors and non-survivors showed a similar pattern to the MIMIC, except for gender. The 16-44 yr and 65-79 yr groups of non-survivors were separately smallest and largest percentages aligned with MIMIC. While the largest percentage agegroup was the 45-64 yr group with 34.5%. The higher APACHE-IVa score was presented for deaths (82.0 points vs. 52.0 points). There was no significant difference in mortality between teaching and non-teaching hospitals.

**Table 1.** Patient characteristics in two study cohorts.

|  | MIMIC (78,550, 11.1% mortality) |  |  |  | eICU-CRD (95,380, 9.0% mortality) |  |  |  |
| --- | --- | --- | --- | --- | --- | --- | --- | --- |
|  | Overall | Survivor | Non-Survivor | *P*-Value | Overall | Survivor | Non-Survivor | *P*-Value |
| Number (%) | 78550 | 69837 (88.9) | 8713 (11.1) |  | 95380 | 86755 (91.0) | 8625 (9.0) |  |
| Age, median [Q1,Q3] | 66.0 [54.0,78.0] | 66.0 [54.0,77.0] | 73.0 [61.0,82.0] | <0.001 | 65.0 [53.0,76.0] | 65.0 [53.0,76.0] | 71.0 [60.0,81.0] | <0.001 |
| Age group (%) |  |  |  |  |  |  |  |  |
| 16-44 | 9806 (12.5) | 9249 (13.2) | 557 (6.4) | <0.001 | 12718 (13.3) | 12151 (14.0) | 567 (6.6) | <0.001 |
| 45-64 | 25942 (33.0) | 23706 (33.9) | 2236 (25.7) |  | 32946 (34.5) | 30567 (35.2) | 2379 (27.6) |  |
| 65-79 | 26341 (33.5) | 23309 (33.4) | 3032 (34.8) |  | 32101 (33.7) | 28813 (33.2) | 3288 (38.1) |  |
| 80- | 16461 (21.0) | 13573 (19.4) | 2888 (33.1) |  | 17615 (18.5) | 15224 (17.5) | 2391 (27.7) |  |
| Gender (%) |  |  |  |  |  |  |  |  |
| Female | 34228 (43.6) | 30238 (43.3) | 3990 (45.8) | <0.001 | 43388 (45.5) | 39460 (45.5) | 3928 (45.5) | 0.927 |
| Male | 44322 (56.4) | 39599 (56.7) | 4723 (54.2) |  | 51992 (54.5) | 47295 (54.5) | 4697 (54.5) |  |
| BMI, median [Q1,Q3] | 27.6 [23.9,32.1] | 27.7 [24.1,32.2] | 26.5 [22.8,31.2] | <0.001 | 27.5 [23.5,32.9] | 27.6 [23.6,33.0] | 26.7 [22.7,32.1] | <0.001 |
| Language group (%) |  |  |  |  |  |  |  |  |
| English | 56659 (72.1) | 51147 (73.2) | 5512 (63.3) | <0.001 | -- | -- | -- | -- |
| Non-English | 21891 (27.9) | 18690 (26.8) | 3201 (36.7) |  | -- | -- | -- | -- |
| Ethnicity (%) |  |  |  |  |  |  |  |  |
| Asian | 2116 (2.7) | 1869 (2.7) | 247 (2.8) | <0.001 | 1398 (1.5) | 1256 (1.4) | 142 (1.6) | 0.001 |
| Black | 7809 (9.9) | 7098 (10.2) | 711 (8.2) |  | 11219 (11.8) | 10301 (11.9) | 918 (10.6) |  |
| Hispanic | 2711 (3.5) | 2504 (3.6) | 207 (2.4) |  | 3659 (3.8) | 3332 (3.8) | 327 (3.8) |  |
| Other | 12001 (15.3) | 10117 (14.5) | 1884 (21.6) |  | 5915 (6.2) | 5418 (6.2) | 497 (5.8) |  |
| White | 53913 (68.6) | 48249 (69.1) | 5664 (65.0) |  | 73189 (76.7) | 66448 (76.6) | 6741 (78.2) |  |
| SOFA, median [Q1,Q3] | 4.0 [2.0,7.0] | 4.0 [2.0,6.0] | 7.0 [5.0,11.0] | <0.001 | 5.0 [3.0,7.0] | 5.0 [3.0,7.0] | 8.0 [5.0,10.0] | <0.001 |
| APACHE-IVa, median [Q1,Q3] | -- | -- | -- | -- | 54.0 [40.0,71.0] | 52.0 [39.0,68.0] | 82.0 [64.0,106.0] | <0.001 |
| CCI score, median [Q1,Q3] | 5.0 [4.0,7.0] | 5.0 [3.0,7.0] | 7.0 [5.0,9.0] | <0.001 | 4.0 [2.0,5.0] | 4.0 [2.0,5.0] | 4.0 [3.0,5.0] | <0.001 |
| Ventilation (%) | 31397 (40.0) | 26357 (37.7) | 5040 (57.8) | <0.001 | 37756 (39.6) | 31831 (36.7) | 5925 (68.7) | <0.001 |
| Code status (%) | 3565 (4.5) | 2323 (3.3) | 1242 (14.3) | <0.001 | 8857 (9.3) | 6785 (7.8) | 2072 (24.0) | <0.001 |
| LOS ICU day, median [Q1,Q3] | 2.4 [1.6,4.5] | 2.3 [1.5,4.1] | 3.9 [2.1,8.0] | <0.001 | 2.3 [1.6,4.0] | 2.2 [1.6,3.9] | 3.6 [2.0,6.8] | <0.001 |
| LOS hospital day, median [Q1,Q3] | 7.5 [4.7,12.9] | 7.5 [4.8,12.7] | 7.5 [3.4,14.8] | <0.001 | 6.2 [3.7,10.6] | 6.2 [3.8,10.4] | 6.0 [3.1,11.8] | <0.001 |
| Pre ICU LOS day, median [Q1,Q3] | 0.1 [0.0,0.7] | 0.1 [0.0,0.7] | 0.1 [0.0,0.8] | <0.001 | 0.2 [0.0,0.4] | 0.2 [0.0,0.4] | 0.1 [0.0,0.7] | <0.001 |

CCI: Charlson Comorbidity Index, LOS: length of stay.

In **Figure 2**, the discriminative comparisons (AUROC) of agegroup, gender, and language with SOFA and APACHE-IVa scores in two study cohorts were presented. Following the previous study, the discriminative performance of AUROC could be categorized into four types poor (<0.7), acceptable (0.7-0.8), excellent (0.8-0.9), and outstanding (>0.9) [1]. The overall performance was 0.744 (0.739-0.75) of SOFA in MIMIC, 0.725 (0.721-0.729) of SOFA in eICU-CRD, 0.822 (0.818-0.828) of APACHE-IVa in eICU-CRD, which was considered to be acceptable or excellent performance in our study cohorts. For agegroup, they all showed a significantly decreased trend with age (*p* < 0.001) in **Figure 2** and **eTable 1** (*‘16-44’, ‘45-64’, ‘65-79’, and ‘80-’:* 0.812, 0.777, 0.732, and 0.705 in MIMIC for SOFA; 0.808, 0.763, 0.706, and 0.673 in eICU-CRD for SOFA; 0.882, 0.839, 0.803, and 0.754 in eICU-CRD for APACHE-IVa). There existed a 1 to 2 level difference in predictive performance between youngest (16-44 yr) and oldest (80-yr) patients. For gender, male’s predictive performance was slightly better than female’s with significant difference (0.748 vs. 0.745; 0.728 vs. 0.721; 0.827 vs. 0.819, *p* value < 0.001). For first language speaking, English speakers’ AUROC was significantly higher than Non-English speakers’ in the MIMIC cohort (0.771 vs. 0.709, *p* value < 0.001). Furthermore, we explored the cross subgroups’ performance of agegroup and gender in the English speaker (Yes or No) group, shown in **eTable 3**. The decreased performance with age was still observed in the English and Non-English group. However, gender performances were not the same as before. In the English speaker group, females’ AUROC was slightly better than males (0.776 vs. 0.771). While females’ were lower than males’ (0.693 vs. 0.721) in the Non-English speaker.

**Figure 2.**
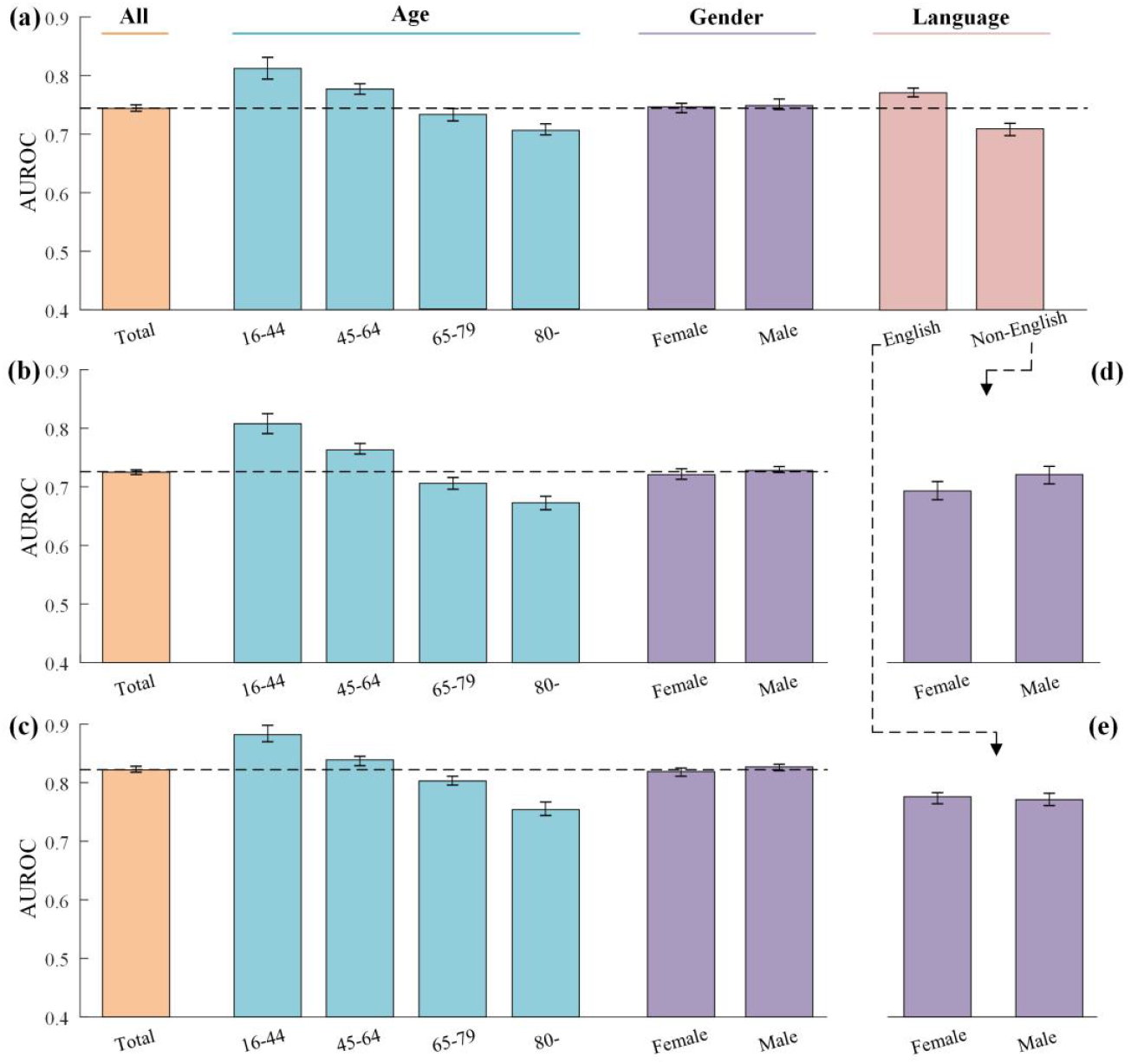
The discriminative comparison of the AUROC for predicting hospital mortality grouped by agegroup, gender, and language. (a) SOFA score in the MIMIC, (b) SOFA score in the eICU-CRD, (c) APACHE-IVa score in the eICU-CRD, (d) and (e) SOFA score in the MIMIC of Non-English and English speakers

The discriminative performance of AUPRC was presented in **eTable 2** and **eTable 3**. The overall performance was 0.304 (0.295-0.312) of SOFA in MIMIC, 0.236 (0.227-0.244) of SOFA in eICU-CRD, 0.382 (0.372-0.397) of APACHE-IVa in eICU-CRD. For agegroup, the performance didn’t show a stable pattern of change with age. The youngest’s was lower than the oldest’s for SOFA score (0.257 vs. 0.356 in MIMIC, 0.21 vs. 0.271 in eICU-CRD). While it was reversed for the APACHE-IVa score (0.411 vs. 0.367). For gender, there was no consistent difference between males and females in the two cohorts. For English proficiency, English speakers’ AUPRC was significantly lower than Non-English speakers’ in the MIMIC cohort (0.304 vs. 0.33, *p* value < 0.001). In the cross subgroup analysis of the MIMIC cohort, there was a consistent difference between the youngest and oldest in both groups (English and Non-English groups). The AUPRCs of gender were the same as before, with no consistent pattern.

In **Figure 3**, the calibration comparisons of SMR in two cohorts by three categories were displayed. As shown in **Figure 3(a)** and **eTable 4**, the fitted mortality function of SOFA, using 10% patients randomly selected, showed a well estimation of the entire cohort’s risk in the MIMIC (8741 death predicted (E) vs. 8713 death observed (O)). While there exists a significant difference in subgroup analysis. For agegroup, SMR increased with age from 0.6 to 1.57. The serious overprediction of the youngest and poor underestimation of the oldest was observed (926 deaths of E vs. 557 deaths of O; 1840 deaths of E vs. 2888 deaths of O). The group of 65-79 yr showed the most accurate prediction (SMR = 1), and the remaining group was kind of overprediction (SMR = 0.76). For gender, the mortality of male patients was overestimated and females’ was underestimated (SMR: 0.92 vs. 1.1). For English proficiency, there exists overprediction for English speakers and serious underestimation for Non-English speakers (6462 deaths of E vs. 5512 deaths of O; 2279 deaths of E vs. 3201 deaths of O). In the eICU-CRD cohort, there exist the same change pattern of SMR by agegroup and gender for SOFA with serious overestimation of youngest and poor underestimation of oldest patients, and more focusing on male patients (**Figure 3(b)**). For the APACHE-IVa score in eICU-CRD cohort, we directly used the mortality estimation function to calculate the risk probabilities, generated by the previous study. There exists an overestimation of hospital mortality for the entire group (12154 deaths of E vs. 8625 deaths of O, SMR 0.71). The serious overestimation was still observed compared with other age groups (SMR: 0.62, 0.69, 0.74, and 0.71). It was more concentrated on females than males (SMR: 0.69 vs. 0.72).

**Figure 3.**
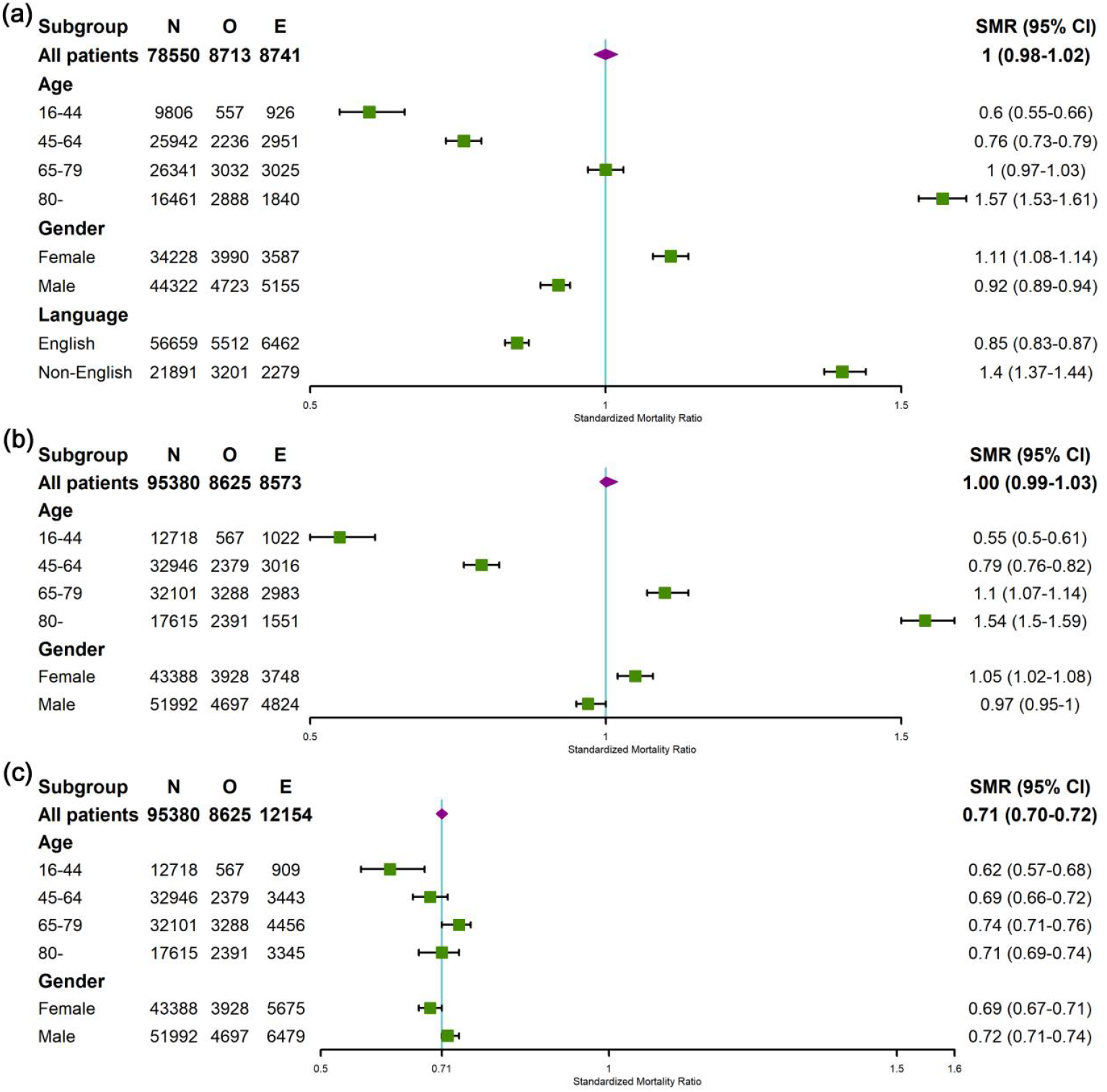
The calibration comparison (SMR) for two clinical scores in the MIMIC and eICU-CRD cohort. (a-b). the SOFA score in two cohorts separately. (c). the APACHE-IVa score in the latter (N: number, O: observation, E: expectation)

The calibration belts of SOFA score by subgroup analysis were shown in **Figure 4**. The 80% and 95 % CI boundaries were both plotted using light and dark grey area respectively in each subplot. The significant deviation from the bisector was tested by the Wald test and the order of polynomial logistic regression was fitted, shown in the top-left corner by P-value and Polynomial degree. The under (overestimation) and over (underestimation) bisector by the deciles of risk were displayed in the lower-right corner. There exist overestimation in the last 6 deciles of risk for the entire cohort (*p* value < 0.001). For agegroup, the youngest group showed an overall overestimation of hospital mortality with the largest confidence interval (Polynomial degree = 2). The 45-64 yr group showed slightly overestimated, and the 65-79 yr group was well-calibrated without significant difference (*p* value =0.392). While the general underestimation was observed in the oldest patients (**Figure 4(b)-(e)**). In **Figure 4(f)** and **(g)**, there exist underestimation in the first 4 deciles and overestimation in the second to last decile for female patients. While the overestimation always exists from the first 3 deciles of risk for males. In **Figure 4(h)** and **(i)**, the general overestimation and large degree of underestimation for English and Non-English speakers were shown. In **eFigure1** and **eFigure2**, the calibration belts of SOFA and APACHE-IVa scores in the eICU-CRD cohort were presented. For the SOFA score, the overestimation in the first 5 deciles for the youngest patients with the largest confidence interval and general underestimation for the oldest patients were obtained. The slight general underestimation and little overestimation in the first decile were observed for females and males, respectively. For the APACHE-IVa score, almost all were overestimated for any subgroups.

**Figure 4.**
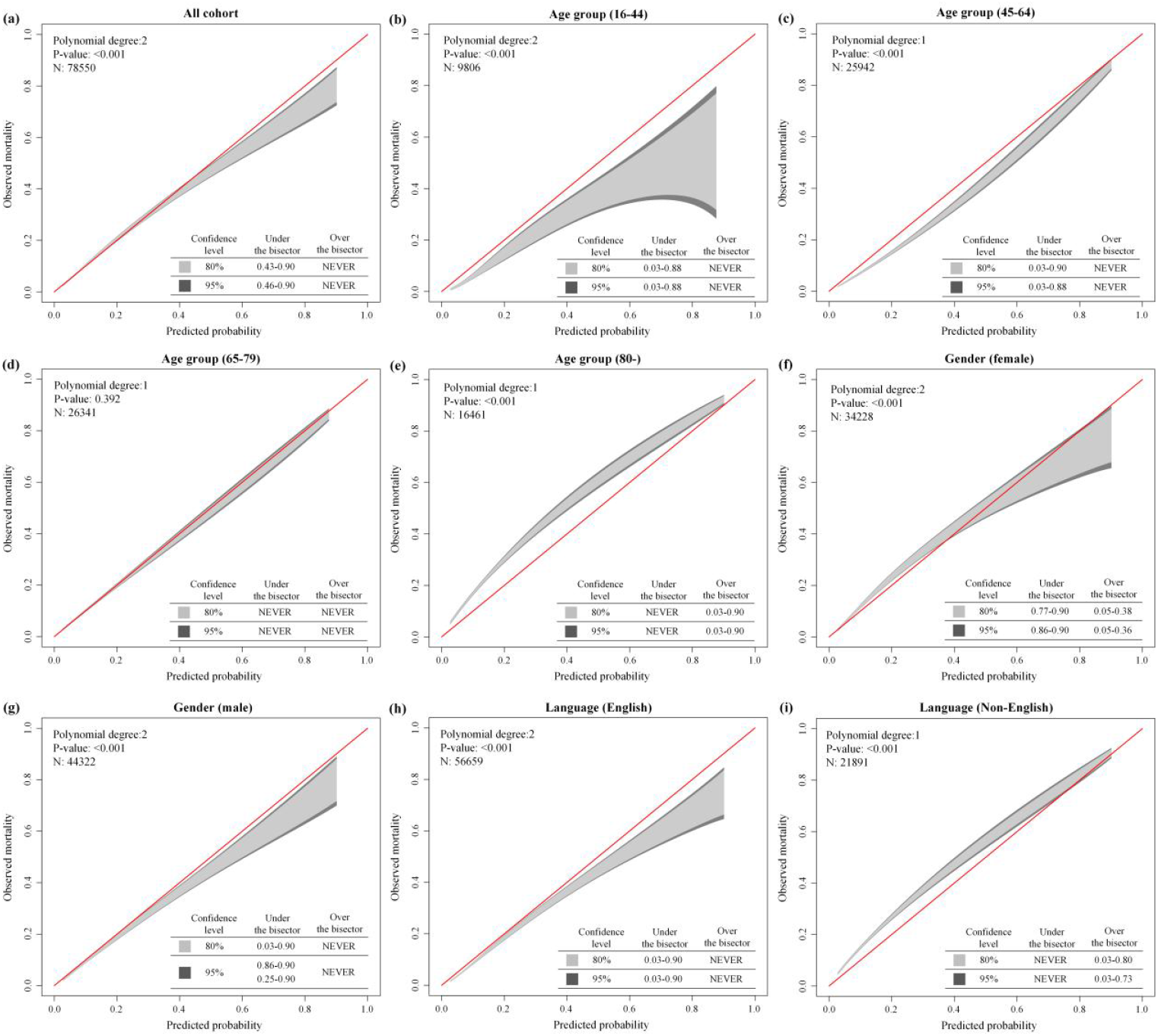
GiViTI calibration belt of SOFA score grouped by age, gender, and language in the MIMIC cohort. (a) total cohort, (b)-(e) four age groups (16-44, 45-64, 65-79, and 80-yr), (f) and (g) gender groups (female and male), (h) and (i) language groups (English speakers and Non-English speakers)

We additionally conducted two types of disease severity categories to comprehensively present the bias of SOFA score on hospital mortality assessment in each subgroup. According to the mortality probability of fitted functions, the prediction and observation of deaths with SMR values in those five risk categories were presented in **eFigure 3** (MIMIC), **eFigure 4** (eICU-CRD), and **eTable 5**. As shown in **eFigure 3(a)** and **(b)**, there exist overall overestimation in the 16-44 and 45-64 yr group, which presented the degree of bias gradually decreased as severity increased. A well consistency of predicting and observing with the 65-79 yr group was observed with somewhat underestimated. For the 80-yr group, a serious ignorance of severity was shown when admitted to ICU with milder diseases. And the underestimation was mitigated with disease severity. In **eFigure 3(c)** and **(d)**, female patients with milder or moderate severity (0∼50%) were underestimated and it was reversed with heavy diseases. While the overpredicted mortality in males was observed regardless of disease severity. In **eFigure 3(e)** and **(f)**, it was obviously overpredicted for English speakers and underestimated for Non-English speakers with milder diseases (SMR: 0.64 vs. 1.73 in 0-5% risk category). The bias degree was mitigated with severity increased. In the eICU-CRD cohort (**eFigure 4**), similar patterns for agegroup were observed except for the 65-79 yr group. No patterns were found for males and females. It seems to be that a higher percentage of women were underestimated and a higher percentage of men were overestimated. In **eTable 6**, we performed the subgroup analysis in all cohorts with three SOFA score categories (0-7, 8-11, over 11). It was still observed the overestimation of youngest and English speaking patients and underestimation of oldest patients in all categories. There exist overprediction of males and underestimation of females when SOFA scores less than 11 points in the MIMIC cohort. While no patterns were observed for gender in the eICU-CRD cohort like the previous result.

The fitted logistic curves of observed mortality with interest scores by agegroup, gender, and language were plotted in our cohorts. In **Figure 5(a)-(c)**, the observed mortality was increased with age when recorded as the same SOFA score in all cohorts. While there was no relationship found for the APACHE-IVa score. In **Figure 5(d)-(f)**, the higher mortality of females was observed with the same SOFA score in all cohorts. But there was still no related finding for the APACHE-IVa score. There exist somewhat higher mortality for Non-English speaking compared with English speaking with the same SOFA score (**Figure 5(g)**). In **eTable 7**, the fitted functions of scores were presented using all patients, respectively. In **eTable 8**, the SOFA scores were adjusted by agegroup with the baseline of the youngest patients. A higher relative risk was observed with increased age in all cohorts, especially the oldest patients. And the *R*^*2*^s were improved when considering the age factor. In **eTable 9**, the SOFA scores were adjusted by gender with the baseline of females. The relatively lower risk was presented for males, especially in the MIMIC cohort. And the *R*^*2*^s were barely changed. In **eTable 10**, Non-English speakers had above 90% higher mortality rate than English speakers when the SOFA score was adjusted by the language variable.

**Figure 5.**
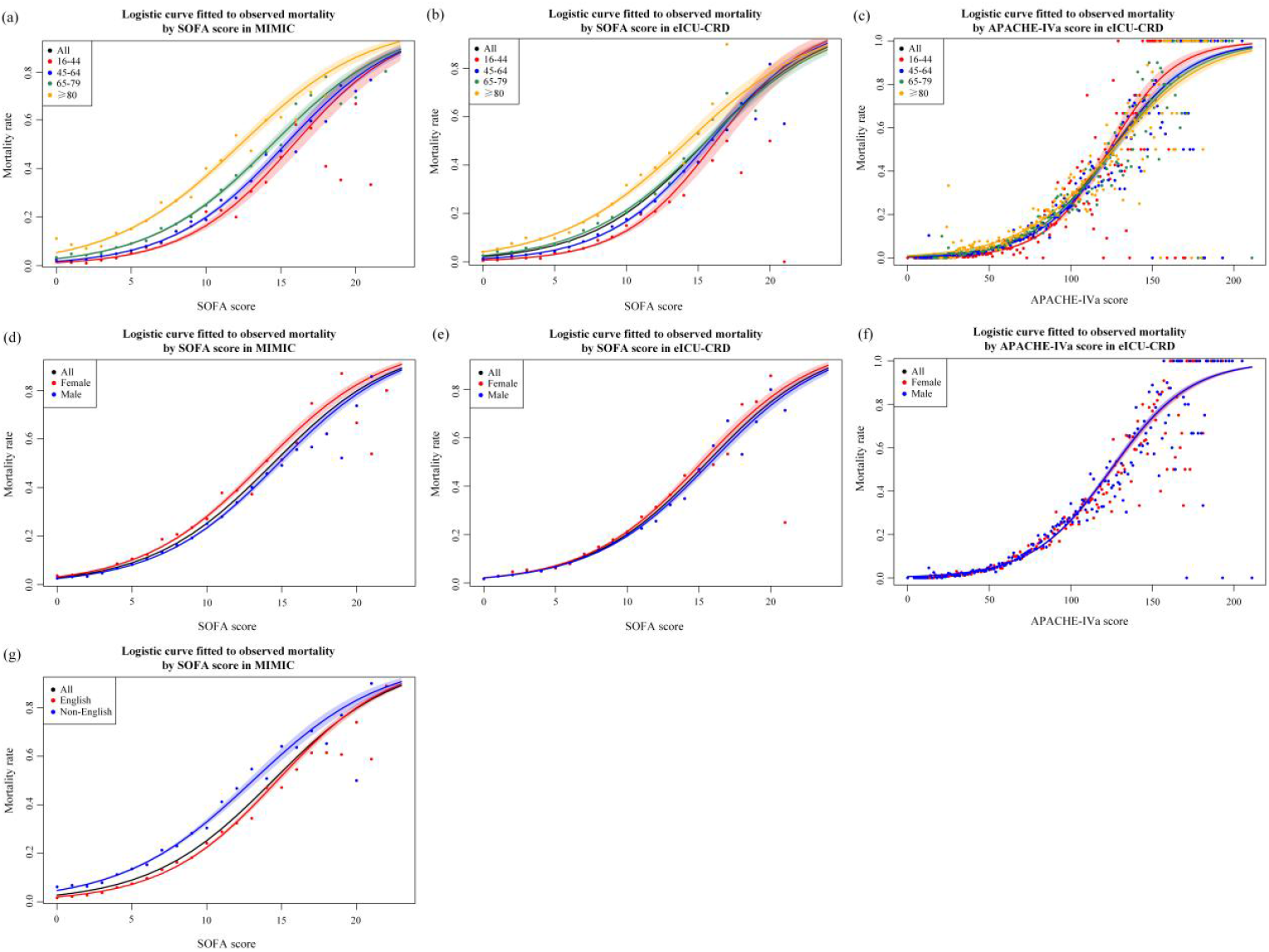
The fitted logistic curve of observed mortality by SOFA and APACHE-IVa score in study cohorts, divided by agegroup, gender, language.

## Discussion

We conducted a retrospective multicenter study to evaluate the performance of ICU prognostic scores covering 189 U.S. hospitals across 19 years stratified by age, gender, and English speaking, respectively. Here present a consistent pattern of SOFA and APACHE-IVa scores in discriminative performance, which was significantly largely decreased from younger to older patients. And slightly better discrimination for males in our study cohorts with significant difference (*p* < 0.001). The poor prediction performance of patients with Non-English speaking was presented when evaluated using the SOFA score. A seriously overpredicted mortality was observed in 16-44 yr group patients for both of them. Uniform standard assessment of SOFA score would lead to serious underestimation for older patients, especially over 80 yrs. Different degrees of calibration bias was also reflected in gender, but two scores didn’t reflect the same concerns. There exist critical underestimation and higher observed mortality of Non-English speaking patients evaluated by SOFA score. The different performance of subgroups and scores remind us that we should be careful with usage and refine evaluation with adjustment, which supports health care systems to adapt to an aging and diverse population and promote the fair clinical practice.

The clinical prognostic scores are often used for severity assessment, patient triage, decision-making, a benchmark of ICU quality, and even for the allocation of scarce treatment resources [7,9]. Notably, developing them has not kept pace with changes in actual population characteristics, especially for widely even daily used SOFA and APACHE-IVa scores. The potential bias and disparities in ethnicity were highly regarded for Black and Hispanic patients in recent [8,9,15,16]. Yet the looming higher percentage of older, vulnerable female and LEP patients face both physical, socioeconomic, and cultural differences as well as communication barriers [17-20]. To our knowledge, there are no studies that large multicenter cohorts were used to analyze the performance and existing bias of those scores in the mentioned groups. It would help us eliminate inherent bias and understand the scope of application to avoid undertreatment with relatively limited clinical resources and investment [17-19]. With the continuous improvement of the modern ICU and the emergence of new treatment technologies, the morbidity and mortality rate of patients is much lower than before [7]. While the rapidly expanding wave of the very old age group (over 80 yrs) brings more challenges and unknowns to intensive care and treatment [17,21-23]. At the same time, physicians also face ethical and practical tests to care for older patients before and during ICU admission [23]. Some studies have found that poorer status, frailty, co-morbidity, and palliative care are associated with worse prognoses in critically ill patients over 80 years old [21,22]. The commonly used SOFA score is an expert consensus, whereas the most up-to-date APACHE-IV score, although taking into account age, was generated based on a population with a median age of 61.5 yrs between 2002 and 2003 [2]. There are still no valid critical illness scores designed for them [17]. Therefore, it is essential and urgent to be informed of the bias introduced by existing systems and tools in the assessment of geriatrics to further guide the design of new scores. In our study, the median age was 66 yrs (11.1% mortality) in MIMIC and 65 yrs (9.0% mortality) in eICU-CRD for the entire group. While the mortality rates were highly increased for the older patients over 80 yrs (33.1% mortality with 21.0% patients in the former, 27.7% mortality with 18.5% patients in the latter). In the discriminative analysis, the performance of the oldest patients appeared a significant decrease compared with the youngest patients for SOFA and APACHE-IVa scores (excellent for the 16-44 yrs group vs. acceptable even poor for the 80-yrs group). In the calibration comparison, we choose a uniform scale to assess disease severity for SOFA score, including fitted function, risk categories, and cutoff categories. The overestimation for youngest patients and underestimation for oldest patients of hospital mortality were observed in the multicenter validations. Although the APACHE-IVa score was adjusted with age, it still showed more attention to the younger group. Here we showed the SMR values with large differences in two cohorts for 16-44 and 80-yrs groups (0.6 vs. 1.57 for SOFA in MIMIC; 0.55 vs. 1.54 for SOFA in eICU-CRD; 0.62 vs. 0.71 for APACHE-IVa). The GiViTI calibration belts also consistently showed the same pattern. Furthermore, there exists underestimated mortality in the 80-yrs group with 0∼50% mortality risk and 0∼11 point SOFA score. The *R*^2^ value became significantly larger when the regression model of the SOFA score incorporated the age variable. The poor discrimination and underestimation of older patients prompt us to discuss and develop a more age-appropriate prognostic system to guide decision-making. However just considering the calibration of age and physiological abnormalities is not enough, since their underlying conditions have changed significantly compared to adults such as medical history, functional status, cognition, co-morbidity, reserve capacity, and frailty degree [17,21-23,41]. These factors need to be considered and quantified to obtain a more precise and useful score for clinicians. It will help physicians make a better evaluation and decide the choice of withholding or withdrawing life-sustaining therapy [23]. And support them to discuss more reasonable treatment options with the patient’s family at an early stage [17,23].

Race and gender disparities in healthcare were persistent hotspots due to biological and social factors [15,18]. Racial inequalities have been given more attention and discussed in recent several years, especially enduring the allocation of scarce healthcare resources and the accuracy of risk severity has been highlighted [8,9,15,16]. Gender bias is more likely to be studied along with race, or to focus more on assessing sepsis and septic shock cohorts [42,43]. However, in the ICU scenarios, the biases using prognostic scores for triage and providing resuscitate treatments have received little attention, and large multicenter studies are needed to evaluate their performance and identify the unconscious bias [15]. Recent studies showed that female patients were more likely to choose limited treatment no matter their disease severity [18]. And females could receive the same level of treatment as males when encountering more serious diseases, especially younger females [18]. As shown in **eTable 11**, in our study, female patients had a somewhat higher admission age in two cohorts (68 vs. 65 yrs in MIMIC, 67 vs. 65 yrs in eICU-CRD) and large ICU admission gaps in the 45-64 yrs group (29.5% vs. 35.8% in MIMIC, 32.0% vs. 36.7% in eICU-CRD). A lower percentage of ventilation received (37.4% vs. 41.9% in MIMIC, 38.8% vs. 40.2 in eICU-CRD) and higher percentage of choosing limitation (5.9% vs. 3.5% in MIMIC, 11.0% vs. 7.9% in eICU-CRD) were also observed. In two cohorts, higher observed mortality of females was presented when measured by SOFA score. Furthermore, we acquired slightly significantly lower discrimination of mortality for females using two score systems (0.745 vs. 0.748 of SOFA in MIMIC, 0.721 vs. 0.728 of SOFA in eICU-CRD, and 0.819 vs. 0.827 of APACHE-IVa in eICU-CRD). While a relatively larger difference in the Non-English group (0.693 vs. 0.721 of SOFA in MIMIC). Despite significant differences in calibration, the two scores didn’t display a consistent pattern. And there were few changes in performance when fitted functions using scores that took gender into account. However, the differences and consistency of prognostic scores, assessed in multicenter cohorts, should deserve our attention.

Limited English proficiency is a common phenomenon in multilingual and multicultural countries in western countries, especially the U.S, and is continuing to have a deep impact on culture, economics, ethics, communication, and healthcare. Language barriers could adversely affect and profoundly challenge many aspects of healthcare covering medical access, quality, comprehension, and satisfaction [44]. Due to communication barriers and inconsistencies in understanding, the dialogue time is prolonged, patients’ medical history cannot be effectively obtained, treatment plans cannot be effectively made, and communication with the patient’s family is affected. These could lead to misdiagnosis, delays in care, reduced quality of care, longer hospital stays, and reduced patient engagement and satisfaction [24,29,44]. In the MIMIC cohort, present in **eTable 12**, we noticed significantly longer ICU and hospital duration for Non-English speaking patients (LOS ICU day: 2.8 vs. 2.3 days; LOS hospital day: 8.0 vs. 7.3 days). Although some studies mentioned no difference in hospital mortality, the Non-English speaking group’s mortality was higher in our population (14.6% vs. 9.7%) [29,44]. LEP is often associated with ethnic minorities, such as Asians, Hispanics, African Americans, etc [24,44]. A number of recent studies have explored the potential discrimination and inherent prejudice against minorities in the process of disease diagnosis and use of aids, as well as the resulting unfair treatment [8,9,15,16]. For the MIMIC cohort, the Asian, Black, and Hispanic separately hold more than 5% but less than 10% of the Non-English population. Furthermore, they were also associated with lower insurance and lower-income. 10.2% of Medicaid (targeting low-income families or people) and 52.5% of Medicare (disabled patients or geriatrics over 65 yrs) for Non-English speakers were contained in our population, and the ratio of choosing limitation treatment was more than 2 times for English-speaking patients [45]. In our evaluation results, the obvious discrimination gap for LEP patients was present using the SOFA score (AUROC: 0.771 vs. 0.709). And the underestimation of mortality (SMR: 0.85 in English vs. 1.4 in non-English) and higher observed mortality rate with the equal SOFA score were acquired for calibration performance (**Figure 3, Figure 4, eFigure 3**, and **Figure 5**). The bias of the Non-English speaking group for prognostic evaluation could further exacerbate disparities and cause irreversible harm to these minority groups.

ICU severity scores are standard and user-friendly tools to quantify patients’ disease severity and perform risk stratification in clinical practice. Many studies have validated the local application of these scores with acceptable performance [14,46]. However, due to the improvement in the medical level and the change in population characteristics, the performance of these scores gradually deteriorated and showed certain bias and unfairness in subgroups. Inaccurate assessments will result in a waste of precious clinical time and resources [7,15]. The researchers suggested updating and adjusting scores before local usage or adding extra variables [13,15]. In our study, the older group showed poor predictive performance and calibration compared to younger patients. Although the APACHE-IVa score has improved the discrimination compared to the SOFA score by incorporating the evaluation of chronic diseases, age, and part of laboratory tests (AUROC of the 80-yr group: 0.673 SOFA vs. 0.754 APACHE-IVa), there is still room for improvement compared to younger patients (AUROC for APACHE-IVa score: 0.882 for the 16-44 yr group vs. 0.754 for the 80-yr group). Since the pre-morbid conditions in geriatrics are more closely associated with prognosis [21,22]. Incorporating assessments of frailty, cognitive function, and daily activities will allow for precise quantification and prediction [21,22]. Of course, this will further increase the clinical workload. But we can try to use electronic medical record systems to automate the extraction and calculation of the information we need or explore more precise biomarkers (it takes more time and money). The lower discrimination and underestimation of female patients were found when using the fitted functions of SOFA score in our research. While the SMR of females was slightly higher than males using the APACHE-IVa score, the discrimination of females was lower for males. So the potential bias, underprediction, for women should be attended to and kept in mind when evaluating the risk of death. The potential disparities of socially vulnerable, LEP patients, were observed using SOFA scores with relatively poor discrimination and underestimation. We calculated there exists a 90% higher mortality rate of Non-English speaking patients compared to English speakers (**eTable 10**). It should raise our high vigilance and conduct more research in other regions to validate if it is reproducible and to what extent.

A clinical score is also an important basis for physicians to implement decision-making including the decision of withholding or withdrawing life support [47]. Underestimation and overestimation of disease severity may lead to patients missing the best opportunity for treatment and time or wasting medical resources [15]. The race has been widely discussed and studied [8,9,15,16]. While the older group actually had a much higher proportion of the 189 hospitals in our study (MIMIC: 54.5% of older patients, 21.0% of oldest patients, 13.4% of Black and Hispanic patients; eICU-CRD: 52.2% of older patients, 18.5% of oldest patients, 15.6% of Black and Hispanic patients). It requires taking into account more than just equity and bias, but also the burden of care, uncertain benefits, and end-of-life practices [23]. However, current guidelines, consensus, and discussion are far from sufficient to alleviate the challenges and pressures faced by clinicians [15,22]. In addition, the COVID-19 pandemic has further exacerbated the need and urgency of research and discussion on this issue due to further scarcity of healthcare resources and higher mortality rates experienced by older patients [48]. End-of-life issues and decisions are a characteristic of critical care medicine [49]. And it was reported that above 15% of ICU patients had a decision on withholding/withdraw life-sustaining treatment (LST) in the U.S. The proportion of decisions is greatly affected by the country and region, which reflect the culture of not avoiding end-of-life decision and the practice of opening interdisciplinary [17,50-52]. Moreover, the judgment of decision to LST is hugely difficult and exists a degree of subjectivity, and its effects might soon be apparent in patients [53]. Since the treatment intensity of geriatrics is often lower than younger patients and variables measured by restrictive therapy could greatly affect the assessment of the acute physiology component of prognostic scoring systems [17,21]. Resuscitation Status was been reported to have an effect on score systems’ performance, especially for evaluating the risk of older patients [17,47]. It can be seen that an accurate assessment of illness severity can help physicians decide whether to adopt restrictive treatments, and the inclusion of considering resuscitation status in the scoring system will enhance their predictive performance. The evidenced-based treatment in cardiovascular exists potential significant health inequalities for females [18,54]. Female patients adopt life-supporting treatments at a lower rate than males and they could receive equal treatments for more serious diseases compared to men [18,55,56]. For the Non-English speaking patients, due to limited ability to communicate and possibly lower-level insurance, their treatment time might be delayed and therapy options might be restricted. For example, targeting the patients admitted to the hospital due to acute stroke, the decision to perform thrombolytic therapy requires, to a large extent, clear information about the time of symptoms onset. While language barriers can have a significant impact on their treatment and outcome [57]. Therefore, analyzing the bias of scoring systems for minority groups, potentially vulnerable, will help us to further improve them, design more equitable assessment systems to support decision-making, and reduce the potential harm to patients due to inherent perceptions.

Another core application of these prognostic scores is to guide patient triage [7,15]. The SOFA score is widely used as a priority score for allocating clinical resources during responses to public health emergencies, COVID-19 pandemic. And the cohorts with a lower SOFA score category, are more likely to receive priority treatment under scarce resources [8,15,16]. However, as viral virulence decreases, mortality decreases, and the crowding out of healthcare resources is largely alleviated [58,59]. We need to formalize the more pressing needs and challenges faced in the ICU, namely how to better assess the prognosis of older patients and provide more effective treatment, and how to provide more equitable access to treatment for other minority groups, not just for ethnicity but also for gender and language. Furthermore, these scores are the benchmark of ICU quality and performance evaluation, which support the promotion of organization management and the development of care quality improvement plans. They are normally used to assess mortality in the overall population as well as subgroups [10,27,60]. Many studies have found that the APACHE-IV, SOFA, and SAPS-III score has good discrimination, but the calibration is poor, and they overestimated the risk of mortality [10,15,60]. Some studies revealed that there were also significant differences in the discrimination of different subgroups, such as patients receiving CABG surgery, and geriatrics. While they were also limited by the sample sizes [11,28]. In our research, the overestimation of the APACHE-IVa score in 208 U.S hospitals was observed (SMR: 0.71 (0.70-0.72)). The different discrimination and calibration across subgroups were discovered between older and younger, female and male, Non-English speakers, and English speakers by analyzing 173,930 patient episodes. Therefore, it’s necessary to assess the bias by subgroup analysis and calibrate the widely used prognostic scores based on locally accumulated data.

Our research has some limitations. First, we excluded patients without both APACHE-IVa and SOFA score records. The missing ratio of APACHE-IVa was actually much higher than the SOFA scores, which lead to a large reduction in our sample size. However, in order to be able to compare the differences between the two scores, the exclusion criteria were adopted. The close population baseline of MIMIC and eICU-CRD also suggested that such exclusion criteria did not materially affect the study results. Second, only a small number of patients in the eICU-CRD dataset have language records that cannot support the study, and about 25.4% of our study episodes in the MIMIC dataset didn’t record the information. We processed the patients with unmarked English as Non-English speakers, which introduced some errors. Because undocumented patients might also communicate in English, the proportion of Non-English speaking patients was higher than actual. Third, we did not exclude patients who choose non-aggressive treatments, which had some impact on the results. However, considering the relatively small proportion (MIMIC 4.5%, eICU-CRD 9.3%), we hope to reflect the actual situation and these patients were retained. We will conduct a more in-depth analysis after removing this part in the future. Fourth, this article only focuses on hospital mortality, and clinicians are also concerned about 30-day and 90-day mortality. The consideration of these long-term outcomes can evaluate the prognosis more comprehensively. While limited by the dataset used, the relevant research cannot be carried out. Fifth, we did not delve into the impact of comorbidity on patient outcomes and study results. In fact, comorbidities are also an important part of affecting patient prognosis. For example, the performance of APACHE-IVa was significantly better than the SOFA score (AUROC of a total cohort in the eICU-CRD: 0.822 (0.818-0.828) vs. 0.725 (0.721-0.729)). Finally, this paper only carried out the analysis in limited U.S. hospitals, while the applicability in Europe or other countries needs to be further studied.

In conclusion, this study comprehensively analyzed the existing bias in the SOFA and APACHE-IVa score across agegroup, gender, and language speaking with attention to discrimination and calibration. Older patients, females, and Non-English speakers were observed with lower discrimination of two scores compared with other corresponding groups. Relative obvious underestimations of mortality risk in older and Non-English speaking groups were noticed. The SOFA and APACHE-IVa showed significant differences in calibration for females and males, but they were not the same pattern. Potential inherent inequities in commonly used prognostic scores could lead to inaccurate assessments and underestimation of illness severity in minority groups, which in turn could delay lifesaving intervention and make inappropriate treatment plans. When using prognostic scores in clinical practice, taking into account the characteristics of these minority groups and targeted measurement variables can promote the performance of predictions and reduce bias and disparities. What is really needed, is to develop more accurate predictive systems for subgroups and to adequately evaluate and calibrate them in actual use scenarios in order to effectively reduce harm to patients and improve the quality of medical care.

## Data Availability

All data produced in the present study are available upon reasonable request to the authors

https://mimic.mit.edu/

https://eicu-crd.mit.edu/

## Author contribution

## Declaration of interest

None declared.

## Supplementary appendix: *Evaluating Prognostic Bias of Critical Illness Severity Scores Based on Age, Gender, and Primary Language in the USA: A Retrospective Multicenter Study*

**eTable 1.** Comparison of AUROC by agegroup, gender, and language in the MIMIC and eICU-CRD cohort.

| Name | Subgroup (*p value*) | AUROC (MIMIC, SOFA) | AUROC (eICU, SOFA) | AUROC (eICU, APACHE-IVa) |
| --- | --- | --- | --- | --- |
| All |  | 0.744 (0.739-0.75) | 0.725 (0.721-0.729) | 0.822 (0.818-0.828) |
| Age group | 16-44 | 0.812 (0.794-0.831) | 0.808 (0.791-0.825) | 0.882 (0.87-0.898) |
|  | 45-64 | 0.777 (0.768-0.786) | 0.763 (0.756-0.774) | 0.839 (0.829-0.845) |
|  | 65-79 | 0.732 (0.721-0.742) | 0.706 (0.696-0.716) | 0.803 (0.796-0.811) |
|  | >=80 | 0.705 (0.697-0.716) | 0.673 (0.661-0.684) | 0.754 (0.744-0.767) |
|  | *P value* | <0.001 | <0.001 | <0.001 |
| Gender group | Female | 0.745 (0.735-0.751) | 0.721 (0.713-0.731) | 0.819 (0.811-0.825) |
|  | Male | 0.748 (0.741-0.759) | 0.728 (0.724-0.734) | 0.827 (0.821-0.832) |
|  | *P value* | <0.001 | <0.001 | <0.001 |
| Language group | English | 0.771 (0.764-0.779) | -- | -- |
|  | Non-English | 0.709 (0.699-0.72) | -- | -- |
|  | *P value* | <0.001 | -- | -- |

**eTable 2.** Comparison of AUPRC by agegroup, gender, and language in the MIMIC and eICU-CRD cohort.

| Name | Subgroup (*p value*) | AUPRC (MIMIC, SOFA) | AUPRC (eICU, SOFA) | AUPRC (eICU, APACHE-IVa) |
| --- | --- | --- | --- | --- |
| All |  | 0.304 (0.295-0.312) | 0.236 (0.227-0.244) | 0.382 (0.372-0.397) |
| Age group | 16-44 | 0.257 (0.224-0.305) | 0.21 (0.181-0.245) | 0.411 (0.366-0.442) |
|  | 45-64 | 0.313 (0.296-0.329) | 0.242 (0.22-0.257) | 0.378 (0.359-0.397) |
|  | 65-79 | 0.312 (0.293-0.326) | 0.243 (0.227-0.258) | 0.395 (0.382-0.414) |
|  | 80- | 0.356 (0.343-0.376) | 0.271 (0.251-0.29) | 0.367 (0.347-0.39) |
|  | *P value* | <0.001 | <0.001 | <0.001 |
| Gender group | Female | 0.314 (0.296-0.323) | 0.235 (0.227-0.248) | 0.382 (0.37-0.393) |
|  | Male | 0.301 (0.287-0.319) | 0.24 (0.228-0.249) | 0.389 (0.369-0.401) |
|  | *P value* | <0.001 | 0.026 | <0.001 |
| Language group | English | 0.304 (0.292-0.312) | -- | -- |
|  | Non-English | 0.33 (0.312-0.347) | -- | -- |
|  | *P value* | <0.001 | -- | -- |

**eTable 3.** Comparison of discriminative performance by cross subgroup in the MIMIC cohort.

| Subgroup | Cross subgroup (*P value*) | AUROC | AUPRC |
| --- | --- | --- | --- |
| English | Age (16-44) | 0.853 (0.826-0.878) | 0.266 (0.229-0.316) |
|  | Age (45-64) | 0.797 (0.786-0.814) | 0.309 (0.286-0.34) |
|  | Age (65-79) | 0.763 (0.752-0.772) | 0.315 (0.297-0.337) |
|  | Age (80- ) | 0.73 (0.719-0.744) | 0.358 (0.337-0.381) |
|  | *P* value | <0.001 | <0.001 |
| Non-English | Age (16-44) | 0.765 (0.724-0.799) | 0.292 (0.235-0.356) |
|  | Age (45-64) | 0.75 (0.732-0.766) | 0.356 (0.315-0.381) |
|  | Age (65-79) | 0.676 (0.657-0.699) | 0.309 (0.283-0.338) |
|  | Age (80- ) | 0.67 (0.653-0.687) | 0.368 (0.338-0.396) |
|  | *P* value | <0.001 | <0.001 |
| English | Gender (Female) | 0.776 (0.764-0.783) | 0.319 (0.297-0.335) |
|  | Gender (Male) | 0.771 (0.761-0.782) | 0.298 (0.282-0.311) |
|  | *P* value | <0.001 | <0.001 |
| Non-English | Gender (Female) | 0.693 (0.678-0.709) | 0.319 (0.297-0.345) |
|  | Gender (Male) | 0.721 (0.705-0.735) | 0.341 (0.315-0.362) |
|  | *P* value | <0.001 | <0.001 |

**eTable 4.**
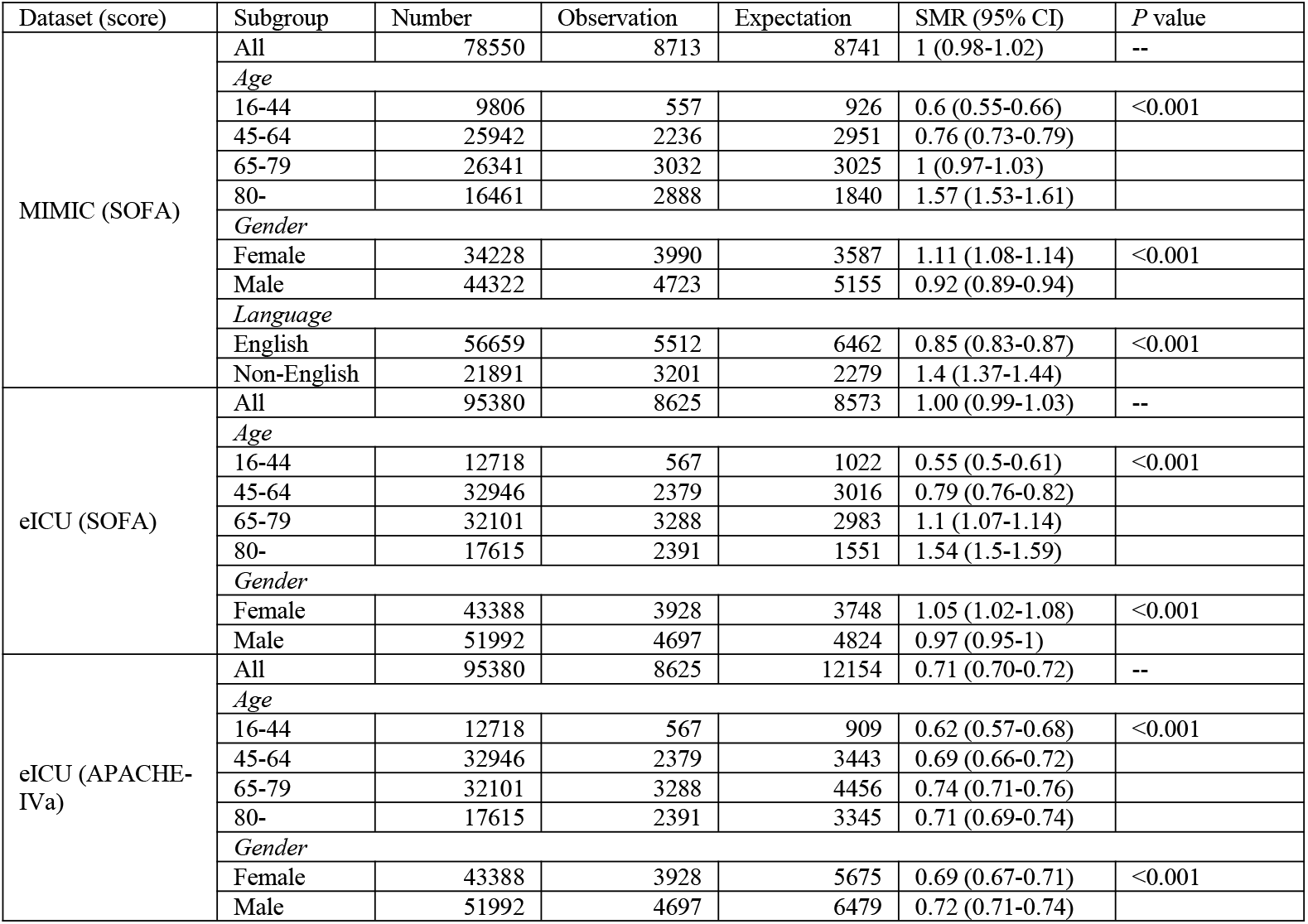
The detailed observation and estimation deaths of SOFA and APACHE-IVa score by age, gender, and language in study cohorts.

**eFigure 1.**
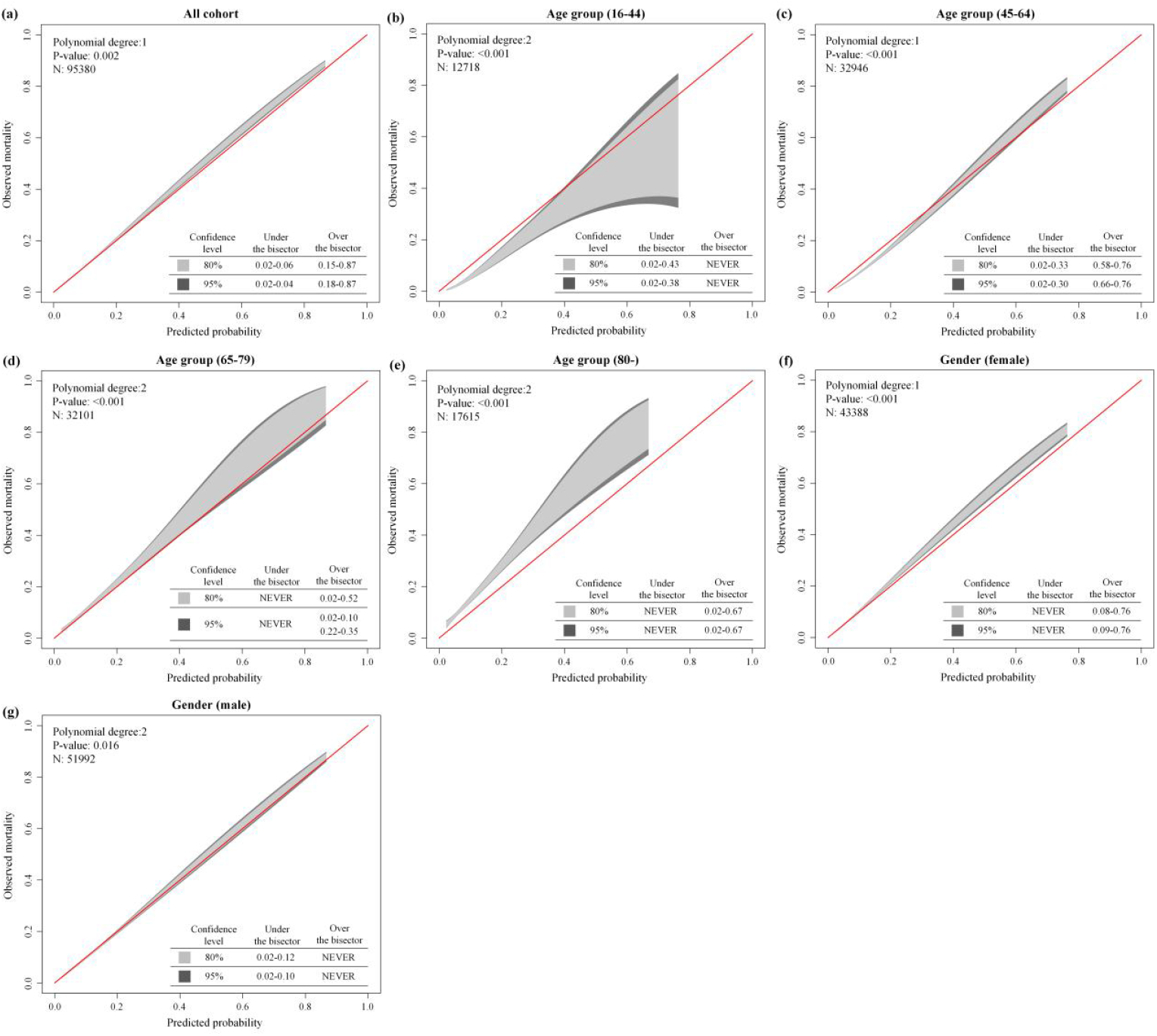
GiViTI calibration belt of SOFA score grouped by age, gender, and language in the eICU-CRD cohort. (a) total cohort, (b)-(e) four age groups (16-44, 45-64, 65-79, and 80- yr), (f) and (g) gender groups (female and male)

**eFigure 2.**
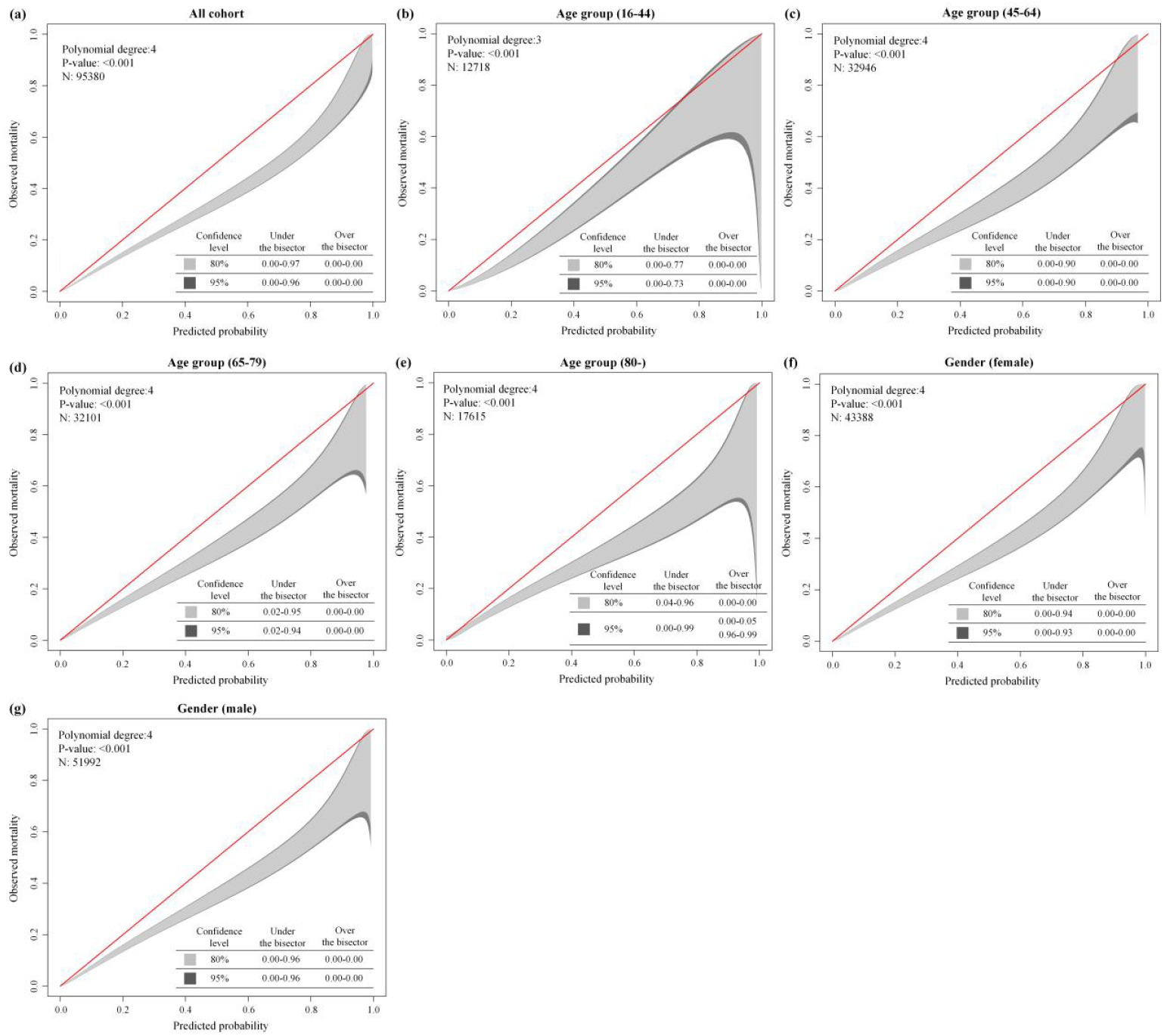
GiViTI calibration belt of APACHE-IVa score grouped by age, gender, and language in the eICU-CRD cohort. (a) total cohort, (b)-(e) four age groups (16-44, 45-64, 65-79, and 80- yr), (f) and (g) gender groups (female and male)

**eFigure 3.**
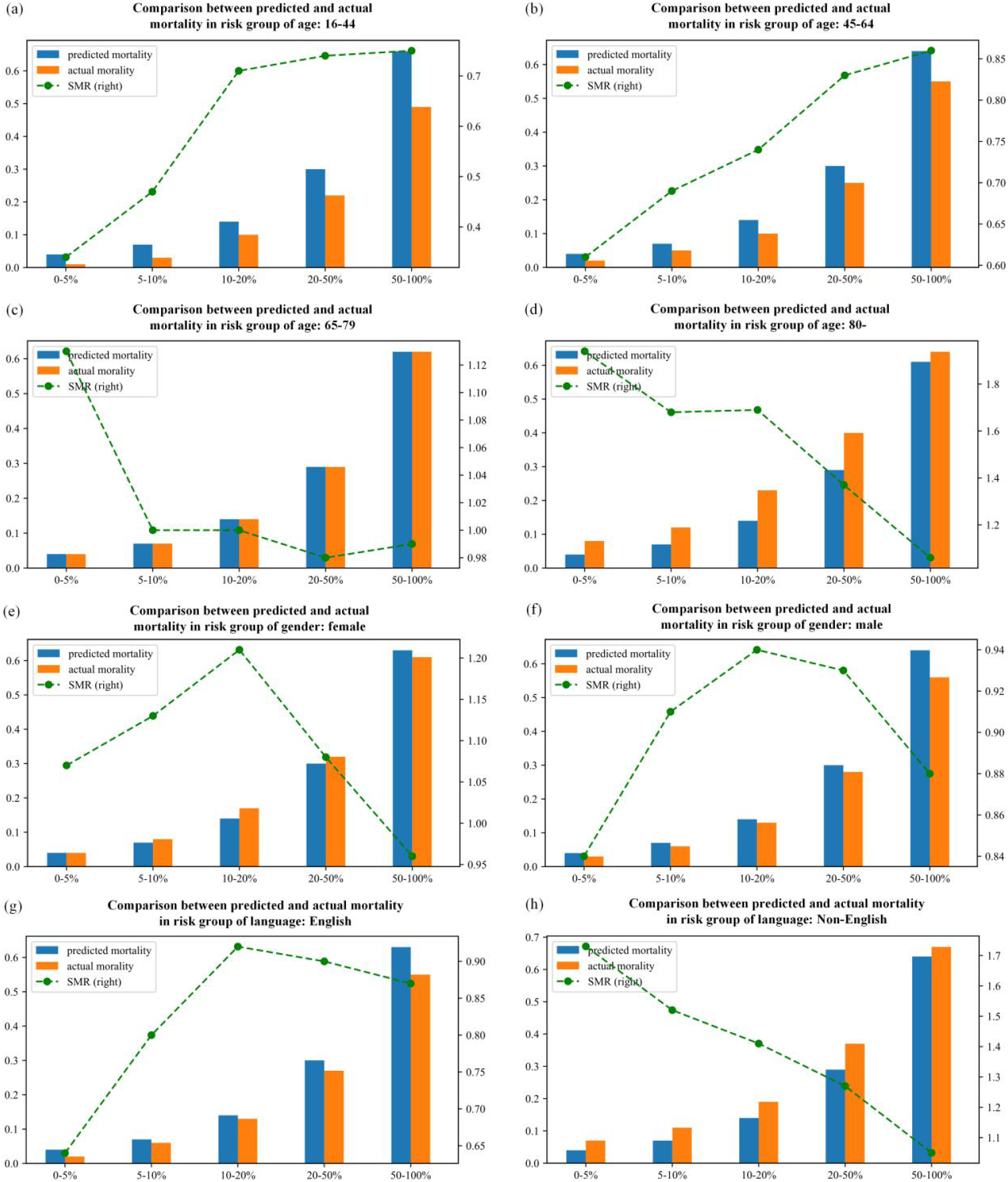
The SOFA score’s SMR trend in five types of risk categories with subgroup analysis in the MIMIC cohort. (a)-(d) for agegroup of 16-44, 45-64, 65-79, and over 80; (e) and (f) for female and male; (g) and (h) for English speakers and Non-English speakers; predicted probability: y-axis on the left; SMR: y-axis on the right

**eFigure 4.**
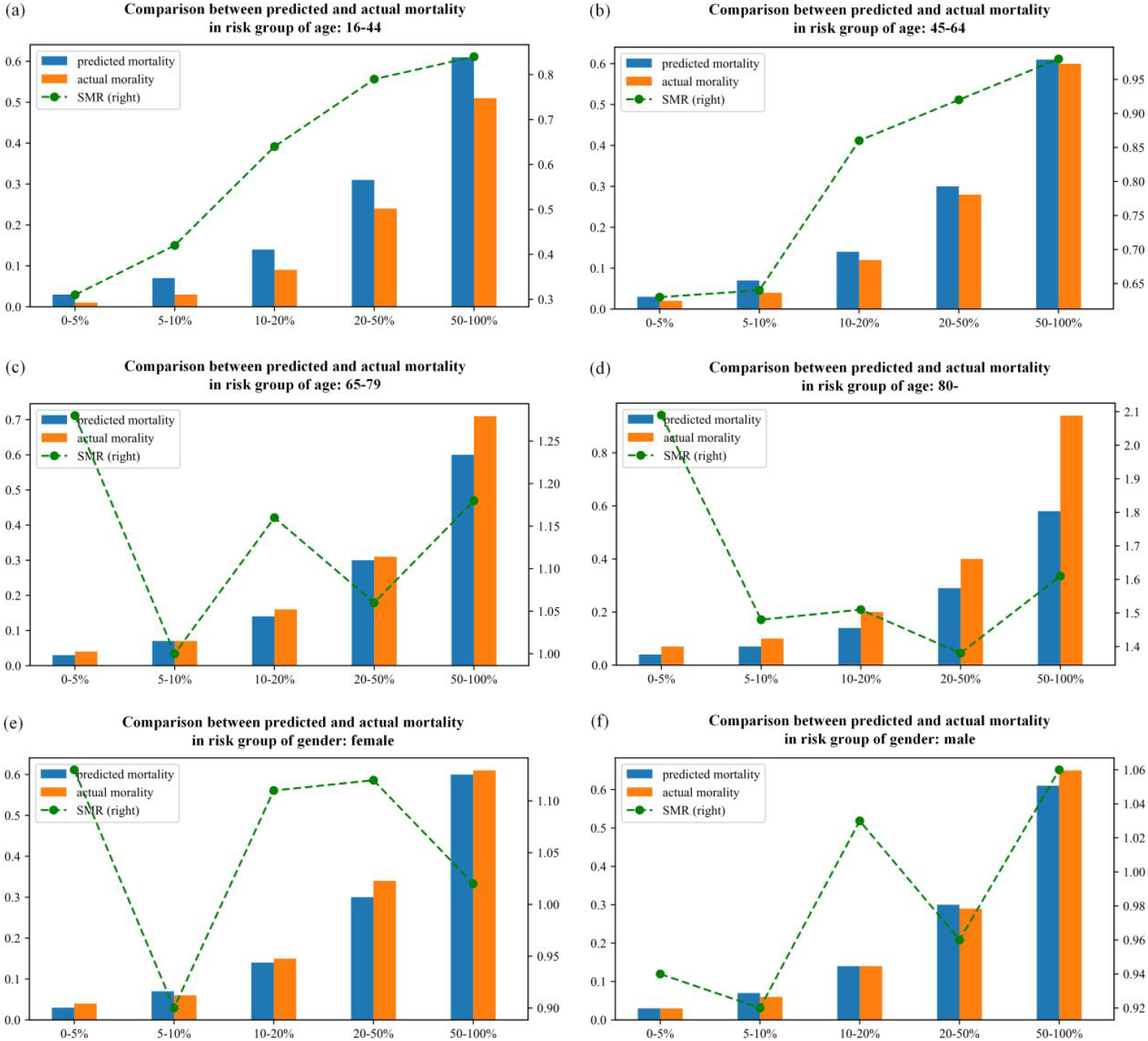
The SOFA score’s SMR trend in five types of risk categories with subgroup analysis in the eICU-CRD cohort. (a)-(d) for agegroup of 16-44, 45-64, 65-79, and over 80; (e) and (f) for female and male; predicted probability: y-axis on the left; SMR: y-axis on the right

**eTable 5.**
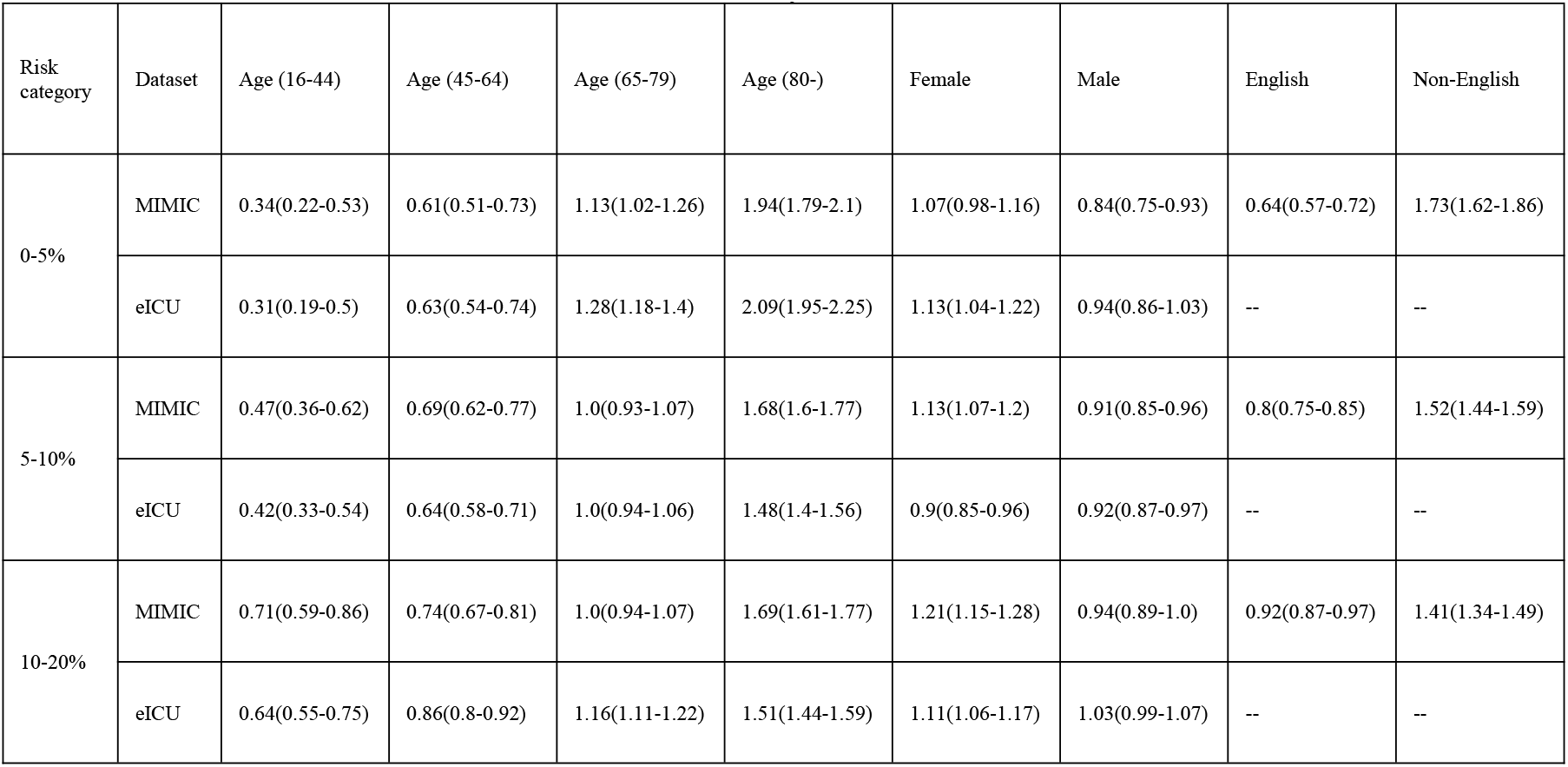
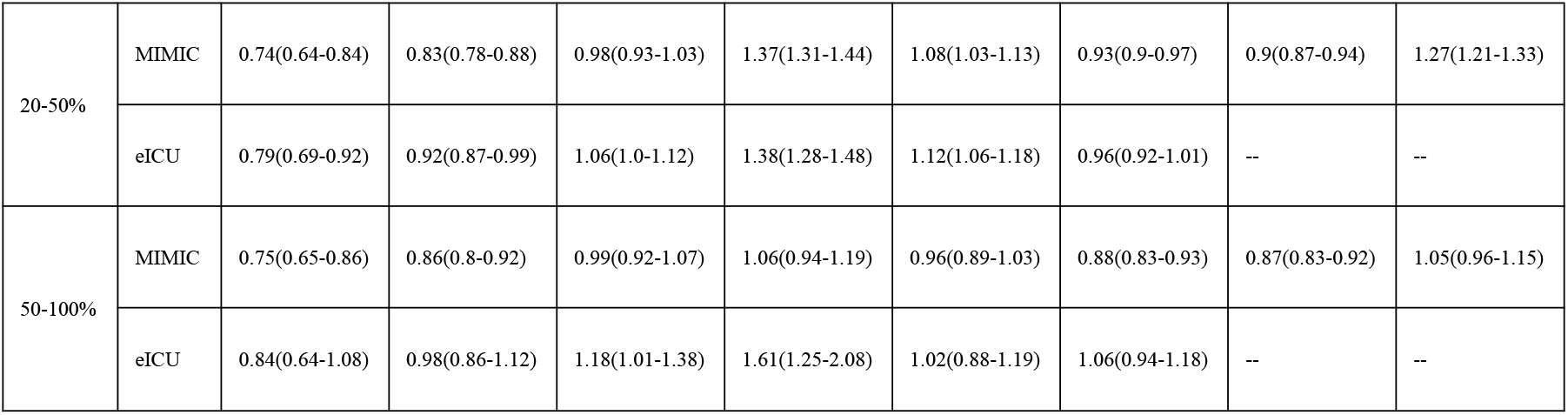
The detailed SMR values across five risk categories in study cohorts by subgroup analysis.

**eTable 6.** The detailed SMR values across three SOFA score categories in study cohorts by subgroup analysis.

| SOFA score | Dataset | Age (16-44) | Age (45-64) | Age (65-79) | Age (80-) | Female | Male | English | Non-English |
| --- | --- | --- | --- | --- | --- | --- | --- | --- | --- |
| 0-7 | MIMIC | 0.48(0.4-0.57) | 0.68(0.63-0.73) | 1.02(0.98-1.07) | 1.75(1.69-1.81) | 1.15(1.1-1.19) | 0.91(0.87-0.95) | 0.8(0.77-0.84) | 1.54(1.48-1.59) |
|  | eICU | 0.4(0.33-0.49) | 0.67(0.62-0.72) | 1.1(1.05-1.15) | 1.61(1.54-1.67) | 0.99(0.95-1.04) | 0.94(0.91-0.98) | -- | -- |
| 8-11 | MIMIC | 0.8(0.69-0.93) | 0.82(0.76-0.89) | 0.98(0.92-1.04) | 1.46(1.39-1.54) | 1.15(1.1-1.21) | 0.93(0.89-0.98) | 0.92(0.88-0.96) | 1.3(1.23-1.37) |
|  | eICU | 0.72(0.62-0.83) | 0.88(0.83-0.94) | 1.12(1.07-1.18) | 1.51(1.43-1.6) | 1.12(1.07-1.18) | 1.02(0.97-1.06) | -- | -- |
| >11 | MIMIC | 0.7(0.62-0.8) | 0.84(0.8-0.89) | 0.99(0.94-1.05) | 1.22(1.14-1.3) | 0.98(0.93-1.03) | 0.91(0.87-0.95) | 0.88(0.85-0.92) | 1.16(1.09-1.23) |
|  | eICU | 0.79(0.68-0.91) | 0.95(0.89-1.02) | 1.07(1.0-1.14) | 1.32(1.21-1.43) | 1.09(1.03-1.16) | 0.98(0.93-1.03) | -- | -- |

**eTable 7.** Logistic regression of SOFA or APACHE-IVa score and hospital death in two cohorts.

|  | Estimate | Std. Error | z value | Pr(>\|z\|) | R<sup>2</sup> |
| --- | --- | --- | --- | --- | --- |
| **MIMIC (SOFA)** |  |  |  |  |  |
| (Intercept) | -3.539531642 | 0.024524367 | -144.3271375 | 0 | 0.1252176 |
| SOFA | 0.245466564 | 0.003073694 | 79.86045123 | 0 |  |
| **eICU (SOFA)** |  |  |  |  |  |
| (Intercept) | -3.843514712 | 0.026640019 | -144.2759757 | 0 | 0.1004896 |
| SOFA | 0.246621933 | 0.003304228 | 74.63829814 | 0 |  |
| **eICU (APACHE-IVa)** |  |  |  |  |  |
| (Intercept) | -5.133000462 | 0.036372562 | -141.1228732 | 0 | 0.1744133 |
| APACHE-IVa | 0.041150103 | 0.000441236 | 93.26093563 | 0 |  |

**eTable 8.** Logistic regression of age and hospital death for SOFA score, with youngest patients (16-44 yr) as baseline group in two cohorts.

|  | Estimate | Std. Error | z value | Pr(>\|z\|) | R<sup>2</sup> |
| --- | --- | --- | --- | --- | --- |
| **MIMIC** |  |  |  |  |  |
| (Intercept) | -4.204521155 | 0.051619647 | -81.45195513 | 0 | 0.1454836 |
| SOFA | 0.251043793 | 0.003146837 | 79.77655101 | 0 |  |
| Age (45-64) | 0.274875021 | 0.05214281 | 5.27158053 | 1.35E-07 |  |
| Age (65-79) | 0.633385127 | 0.050756517 | 12.47889265 | 9.73E-36 |  |
| Age (80-) | 1.218415693 | 0.051356789 | 23.72453011 | 2.01E-124 |  |
| **eICU** |  |  |  |  |  |
| (Intercept) | -4.563560815 | 0.051110477 | -89.28816796 | 0 | 0.1170831 |
| SOFA | 0.251726502 | 0.003378923 | 74.4990323 | 0 |  |
| Age (45-64) | 0.390021198 | 0.049759972 | 7.83805096 | 4.58E-15 |  |
| Age (65-79) | 0.791323656 | 0.04849145 | 16.31882855 | 7.25E-60 |  |
| Age (80-) | 1.204167047 | 0.050148322 | 24.01211058 | 2.08E-127 |  |

**eTable 9.** Logistic regression of gender and hospital death for SOFA score, with female patients as baseline group in two cohorts.

|  | Estimate | Std. Error | z value | Pr(>\|z\|) | R<sup>2</sup> |
| --- | --- | --- | --- | --- | --- |
| **MIMIC** |  |  |  |  |  |
| (Intercept) | -3.416394609 | 0.027034176 | -126.3731733 | 0 | 0.1270618 |
| SOFA | 0.247697487 | 0.003087335 | 80.23019534 | 0 |  |
| Male | -0.245458844 | 0.024392986 | -10.06268135 | 8.08E-24 |  |
| **eICU** |  |  |  |  |  |
| (Intercept) | -3.797982778 | 0.02897362 | -131.0841641 | 0 | 0.1007511 |
| SOFA | 0.247355802 | 0.003311635 | 74.6929634 | 0 |  |
| Male | -0.092211735 | 0.023679052 | -3.894232484 | 9.85E-05 |  |

**eTable 10.** Logistic regression of language and hospital death for SOFA score, with English speakers as baseline group in two cohorts.

|  | Estimate | Std. Error | z value | Pr(>\|z\|) | R<sup>2</sup> |
| --- | --- | --- | --- | --- | --- |
| (Intercept) | -3.789315087 | 0.027487162 | -137.8576329 | 0 | 0.1362904 |
| SOFA | 0.252710612 | 0.003125603 | 80.85179836 | 0 |  |
| Non-English | 0.641189398 | 0.025632063 | 25.01513014 | 4.19E-138 |  |

**eTable 11.**
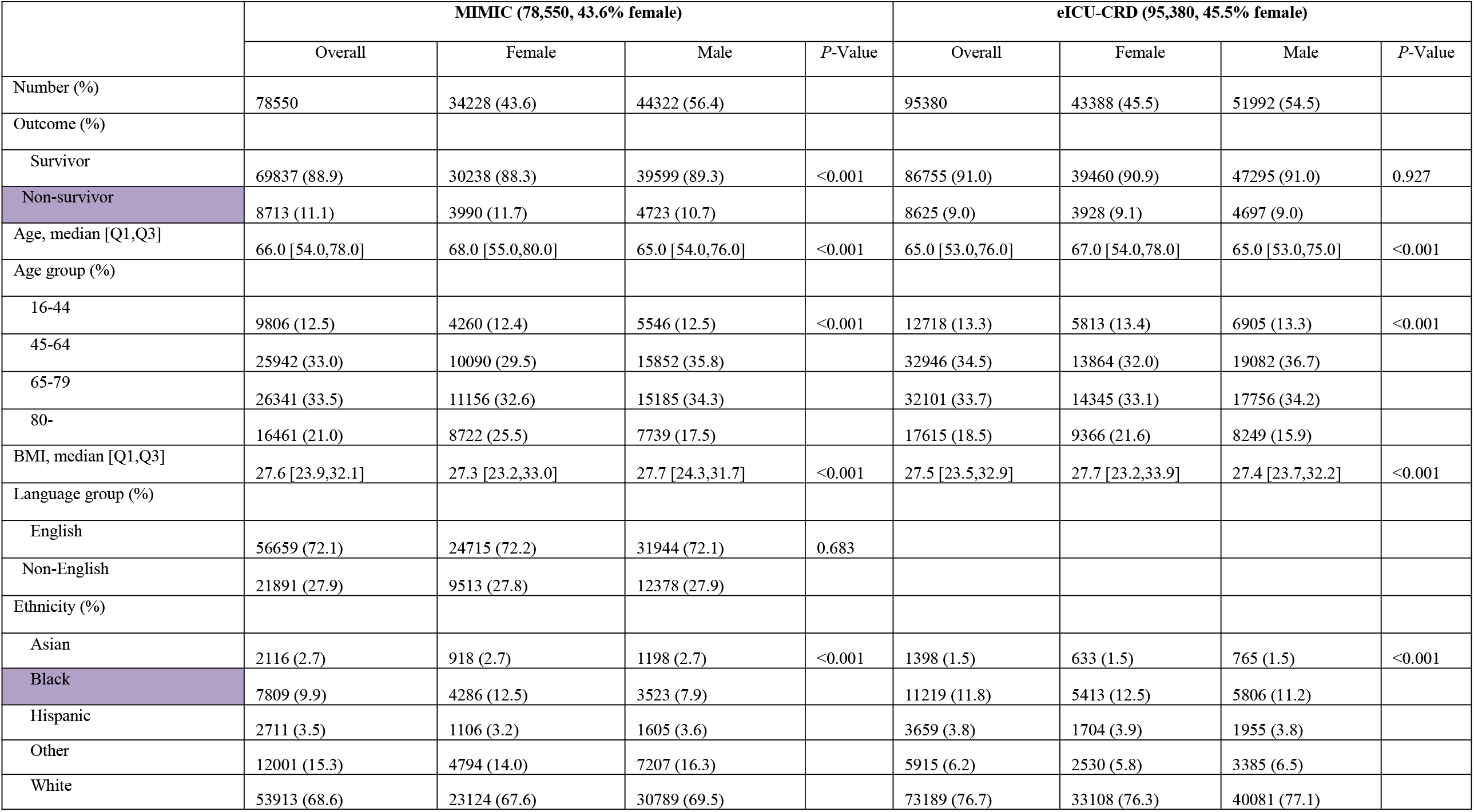
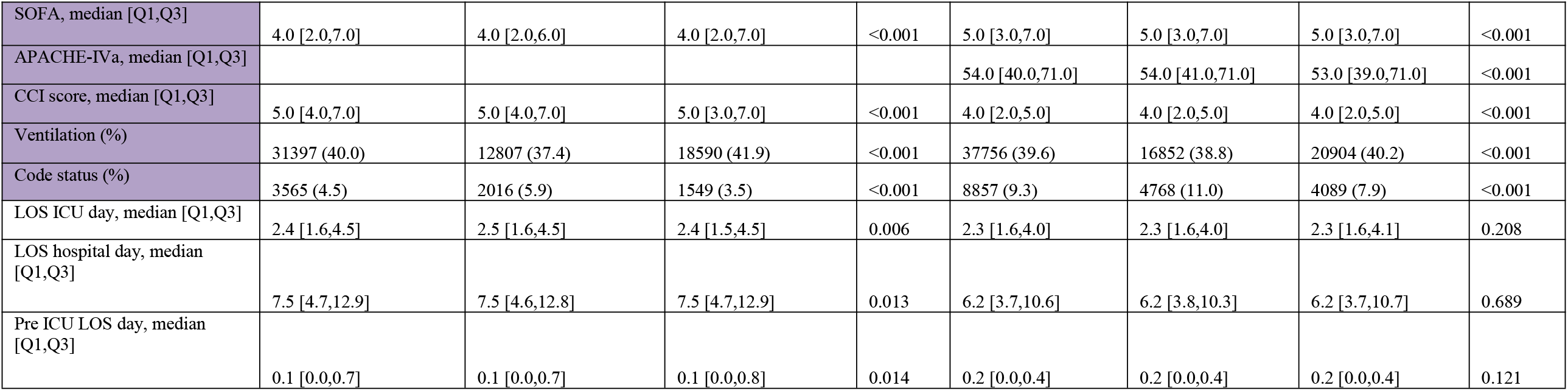
Patient characteristics grouped by gender in two study cohorts.

**eTable 12.**
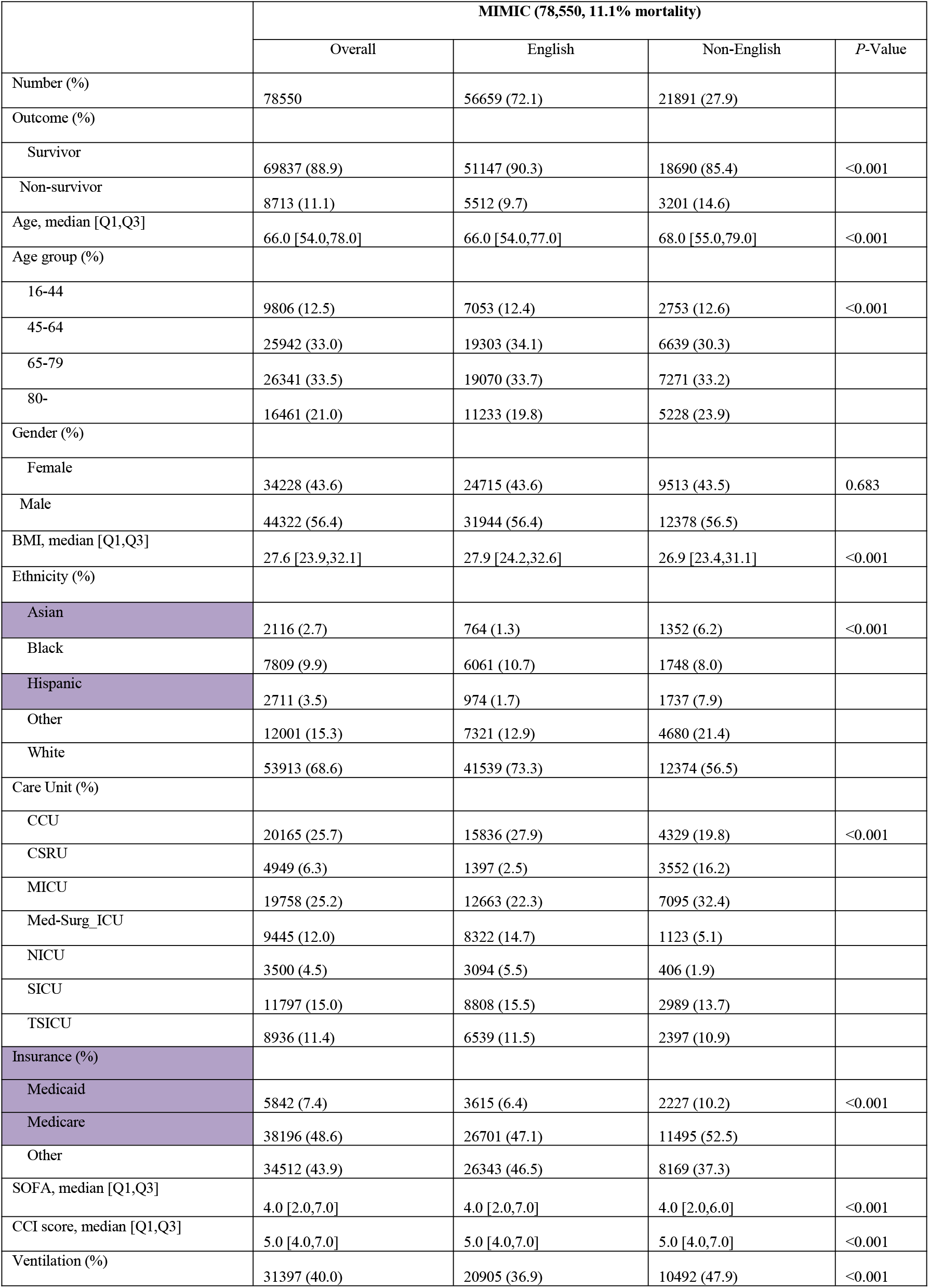
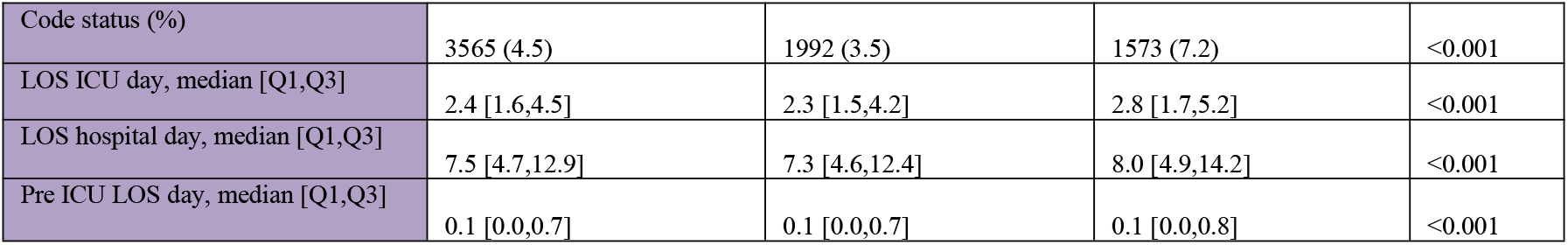
Patient characteristics grouped by language in MIMIC.

|  | MIMIC (78,550, 11.1% mortality) |  |  |  |
| --- | --- | --- | --- | --- |
|  | Overall | English | Non-English | P-Value |
| Number (%) | 78550 | 56659 (72.1) | 21891 (27.9) |  |
| Outcome (%) |  |  |  |  |
| Survivor | 69837 (88.9) | 51147 (90.3) | 18690 (85.4) | <0.001 |
| Non-survivor | 8713 (11.1) | 5512 (9.7) | 3201 (14.6) |  |
| Age, median [Q1,Q3] | 66.0 [54.0,78.0] | 66.0 [54.0,77.0] | 68.0 [55.0,79.0] | <0.001 |
| Age group (%) |  |  |  |  |
| 16-44 | 9806 (12.5) | 7053 (12.4) | 2753 (12.6) | <0.001 |
| 45-64 | 25942 (33.0) | 19303 (34.1) | 6639 (30.3) |  |
| 65-79 | 26341 (33.5) | 19070 (33.7) | 7271 (33.2) |  |
| 80- | 16461 (21.0) | 11233 (19.8) | 5228 (23.9) |  |
| Gender (%) |  |  |  |  |
| Female | 34228 (43.6) | 24715 (43.6) | 9513 (43.5) | 0.683 |
| Male | 44322 (56.4) | 31944 (56.4) | 12378 (56.5) |  |
| BMI, median [Q1,Q3] | 27.6 [23.9,32.1] | 27.9 [24.2,32.6] | 26.9 [23.4,31.1] | <0.001 |
| Ethnicity (%) |  |  |  |  |
| Asian | 2116 (2.7) | 764 (1.3) | 1352 (6.2) | <0.001 |
| Black | 7809 (9.9) | 6061 (10.7) | 1748 (8.0) |  |
| Hispanic | 2711 (3.5) | 974 (1.7) | 1737 (7.9) |  |
| Other | 12001 (15.3) | 7321 (12.9) | 4680 (21.4) |  |
| White | 53913 (68.6) | 41539 (73.3) | 12374 (56.5) |  |
| Care Unit (%) |  |  |  |  |
| CCU | 20165 (25.7) | 15836 (27.9) | 4329 (19.8) | <0.001 |
| CSRU | 4949 (6.3) | 1397 (2.5) | 3552 (16.2) |  |
| MICU | 19758 (25.2) | 12663 (22.3) | 7095 (32.4) |  |
| Med-Surg_ICU | 9445 (12.0) | 8322 (14.7) | 1123 (5.1) |  |
| NICU | 3500 (4.5) | 3094 (5.5) | 406 (1.9) |  |
| SICU | 11797 (15.0) | 8808 (15.5) | 2989 (13.7) |  |
| TSICU | 8936 (11.4) | 6539 (11.5) | 2397 (10.9) |  |
| Insurance (%) |  |  |  |  |
| Medicaid | 5842 (7.4) | 3615 (6.4) | 2227 (10.2) | <0.001 |
| Medicare | 38196 (48.6) | 26701 (47.1) | 11495 (52.5) |  |
| Other | 34512 (43.9) | 26343 (46.5) | 8169 (37.3) |  |
| SOFA, median [Q1,Q3] | 4.0 [2.0,7.0] | 4.0 [2.0,7.0] | 4.0 [2.0,6.0] | <0.001 |
| CCI score, median [Q1,Q3] | 5.0 [4.0,7.0] | 5.0 [4.0,7.0] | 5.0 [4.0,7.0] | <0.001 |
| Ventilation (%) | 31397 (40.0) | 20905 (36.9) | 10492 (47.9) | <0.001 |
| Code status (%) | 3565 (4.5) | 1992 (3.5) | 1573 (7.2) | <0.001 |
| LOS ICU day, median [Q1,Q3] | 2.4 [1.6,4.5] | 2.3 [1.5,4.2] | 2.8 [1.7,5.2] | <0.001 |
| LOS hospital day, median [Q1,Q3] | 7.5 [4.7,12.9] | 7.3 [4.6,12.4] | 8.0 [4.9,14.2] | <0.001 |
| Pre ICU LOS day, median [Q1,Q3] | 0.1 [0.0,0.7] | 0.1 [0.0,0.7] | 0.1 [0.0,0.8] | <0.001 |

## Notes

### Competing Interest Statement

The authors have declared no competing interest.

### Summary of Updates

author affiliations updated

